# A Randomized Controlled Trial Comparing Soy-Pea Protein to Animal Protein in Adults with Crohn’s Disease

**DOI:** 10.64898/2026.05.20.26353678

**Authors:** Abigail Raffner Basson, Jeffry Katz, Vu Nguyen, Drishtant Singh, Paola Menghini, Adrian Gomez-Nguyen, Jennifer Sieg, Madison Bell, Kavitha Thamma, Gina Ponzani, Abdullah Osme, Alexander Rodriguez-Palacios, Fabio Cominelli

## Abstract

**Background and Aims:** Diet plays a critical role in managing Crohn’s disease (CD) inflammation. We assessed whether dietary replacement of animal protein (AnimalP) by soy-pea protein (SoyP) decreases the pro-inflammatory potential of gut microbiota and intestinal inflammation in CD patients.

**Design:** In an open-label, randomized controlled feeding trial at University Hospitals Cleveland Medical Center, CD participants and healthy controls were randomized (1:1) to a soy-pea or animal protein diet for 7-days. Primary outcomes were the absolute difference (Δd7-d0) in; Crohn’s Disease Activity Index (CDAI) score and fecal myeloperoxidase (MPO). Secondary outcomes included fecal calprotectin (FC) and high-sensitivity C-reactive protein (hsCRP). Murine fecal transplantation experiments were performed to determine the inflammatory potential of diet-altered gut microbiota.

**Results:** The study randomized 66 participants, of whom 60 were included in the final analysis (n=31 CD, n=29 HC). After 7 days, CD participants assigned to the SoyP diet were more likely than those assigned to the AnimalP diet to demonstrate reductions in fecal MPO (RR=2.30, 95% CI: 1.04-4.85, P=0.032) and HBI (RR=4.68, 95% CI: 1.22-17.98, P=0.009). The association between SoyP and reduced fecal MPO remained significant after Bonferroni adjustment (FWER-adjusted P=0.006). A similar directional trend was observed for CDAI (RR=1.52, 95% CI: 0.89–2.58, P=0.135), although this did not reach statistical significance. No participants experienced worsening of CDAI. The exploratory post hoc rank-based CDAI-MPO composite score was lower in the CD-SoyP versus CD-AnimalP group (median [IQR]: 5 [4-6] vs 8 [7-9]; P=0.012). Stratified analyses demonstrated significant reductions in fecal MPO among CD participants with lower baseline disease activity (CDAI <150; P<0.0001), but not among those with higher disease activity (P=0.799).

**Conclusion:** Short-term addition of plant-based soy-pea protein within a controlled diet was associated with reductions in objective inflammatory markers in CD, with evidence of greater effects among participants with lower baseline disease activity. ClinicalTrials.gov, Number NCT04065048.

## INTRODUCTION

The inflammatory bowel disease (IBD) subtype Crohn’s disease (CD) is an incurable disorder of the gastrointestinal tract that follows periods of relapse (active disease) and clinical remission. Current therapies aim to control intestinal inflammation, but not all patients respond, and up to 50% lose response over time^1^. Moreover, a significant proportion of patients continue to exhibit residual subclinical inflammation despite symptom control^2, 3^. This persistent inflammatory burden is associated with subsequent disease relapse and loss of therapeutic response^4^. Therefore, alternative therapies that can help alter the natural disease progression while featuring a safe side effect profile are needed for patients with CD.

Dietary interventions are a unique opportunity to improve CD outcomes and could serve as strong drivers to promote or suppress inflammation. The potential benefit of diet in CD is exemplified by the high remission rate observed with exclusive enteral nutrition (EEN) in pediatric CD patients^5^. Therapeutic diets such as the Specific Carbohydrate Diet (SCD), the Mediterranean Diet, and the Crohn’s Disease Exclusion Diet (CDED) have also shown benefit in modulating inflammation in CD^6-8^, although individual responses vary, and not all individuals are able to maintain long-term adherence. There is also evidence that dietary modifications, including intake of anti-inflammatory foods, are associated with metabolic and microbial changes in patients with ulcerative colitis in clinical remission that are linked to reduced subclinical inflammation^9^. As such, there remains a need to explore additional, potentially simpler dietary strategies that may be effective, at least in a subset of patients.

Using animal models, we recently showed that replacing animal protein with soy and pea protein in the context of an American (‘Western’) diet significantly reduced intestinal inflammation in multiple murine models of experimental IBD^10^. Specifically, the soy-pea-based diet led to marked reductions in fecal myeloperoxidase (MPO) activity, histological inflammation, as well as improvements in gut barrier integrity, irrespective of the baseline microbiota composition^10^. These findings suggested that protein source alone, without altering other macronutrient components, can meaningfully influence inflammatory outcomes. Importantly, survey-based data from our patient population indicate that individuals with Crohn’s disease consume soy-based foods significantly less frequently than healthy controls^11^, highlighting a modifiable dietary gap and a clinically actionable target. Together with our preclinical findings and supporting association studies^12^, these data provide a strong rationale to directly test the effects of soy-pea protein, a plant-based dietary protein source, in humans with CD.

Here we present the result of an open-label 7-day randomized controlled trial (RCT) evaluating whether the replacement of animal protein by soy-pea protein in a controlled diet reduces the pro-inflammatory potential of gut microbiota and intestinal inflammation in patients with CD. Healthy controls (HC) were included to contextualize biomarker changes and assess the safety of the dietary intervention. To understand the role of microbiome changes on inflammation, we performed fecal microbiota transplantation (FMT) of diet-altered human microbiomes into germ-free (GF) mice treated with dextran sodium sulfate (DSS). We hypothesized that replacing animal protein with soy-pea protein would reduce intestinal inflammation and improve symptoms in CD, be safe and well-tolerated in both CD and HC participants, and induce anti-inflammatory microbiota shifts capable of attenuating colitis in mice. Overall, we show that short-term addition of plant-based soy-pea protein was associated with improvements in symptom-based indices and objective inflammatory markers, with the greatest impact seen in patients with lower baseline disease activity.

## METHODS

### Study Setting and Participants

This open-label RCT compared the effects of two identical diets differing only in the type of dietary protein, a soy-pea protein-based diet (SoyP) vs an animal protein-based diet (AnimalP), consumed over 7 consecutive days in male and female patients with CD, or healthy controls (HC). Between November 2019 to November 2022, CD outpatients were recruited during routine visits at the Digestive Health Institute (DHI) at University Hospitals Cleveland Medical Center (UHCMC). Healthy controls were recruited from the same population giving rise to the CD participants. The Patient and Public Involvement Statement as well as a detailed description of the study methods are provided in the **Supplementary Materials**.

### Inclusion and Exclusion Criteria

Adult patients (aged 18-65 years) with CD established by standard endoscopic, histologic and radiologic criteria, able to take oral nutrition, and with internet access were included. Harvey Bradshaw Index (HBI) score of <4 were considered ‘CD remission’, scores from 5 to 7 considered ‘mild disease’ and scores of >8 considered ‘CD moderate disease’. The HBI was used pragmatically during screening and recruitment because it provided an efficient and feasible approach for clinical assessment of disease activity in the outpatient setting.

Exclusion criteria were pregnancy/lactation, short bowel syndrome, hospitalized patients, body mass index <19kg/m^2^ or >45kg/m^2^, individuals who lack consent capacity, clinically significant liver/gallbladder/pancreatic dysfunction, uncontrolled diabetes, known drug abuse, known parasitic disease of digestive system, ostomy, known symptomatic intestinal stricture, concurrent malignancy, soy/nut/gluten allergy, currently consuming a soy-based diet (determined by 24-hour recall at enrollment), documented *Clostridium difficile* infection within 4 weeks of screening, start of new, or changes to existing IBD-related medications within 8 weeks of screening, history of <3 bowel movements per week.

### Randomization

Participants with CD and HC were randomized separately in a 1:1 ratio using previously generated random blocks of 2 and 4 created in Microsoft Excel, with assignments placed in sequentially numbered, sealed, opaque envelopes. Envelopes were used consecutively and opened only after informed consent was obtained and patients’ agreement to the randomization process. Study investigators were blinded to the group assignment sequence.

### Study Foods

All meals and snacks (breakfast, lunch, dinner, and two snacks) were provided (Bionutrition, Dahms Clinical Research Unit, UHCMC). Participants were instructed to consume only the study-provided foods and received dietary counseling from a dietitian, including guidance on food preparation. No restrictions were placed on meal timing. Diets were preceded by a 12-hr overnight fast and were immediately followed by the return to usual eating habits after day 7.

### Recruitment

Participants were identified using appointment lists from the DHI at UHCMC and the UH EMR (TriNetX). Recruitment letters were sent, followed by telephone contact, and hospital advertisement flyers were used. Participants were given the option to participate in the study for a second time and consume the other diet not originally assigned (6-week wash out).

### Outcome Measures

Primary and key secondary outcomes were measured at baseline (day 0; d0) and at day 7 (d7) of the study intervention. Herein, our study prospectively specified two primary outcomes representing complementary aspects of disease response: 1) absolute difference (d7-d0) in CDAI as a patient-reported symptom-based measure, and 2) absolute difference (d7-d0) in fecal MPO as an objective inflammatory biomarker. The CDAI was selected because it is a widely used standardized outcome measure in IBD-diet clinical trials^6, 13^ and incorporates multiple disease-related variables beyond patient-reported symptoms alone, including hematocrit, use of antidiarrhea medications, body weight, abdominal mass on examination, and extraintestinal manifestations. CDAI score of <150 considered remission^14^. Baseline CDAI score calculation was based on symptom data prior to initiation of the dietary intervention (d0), whereas the study day 7 CDAI score incorporated symptom data collected during the 7-day intervention period. The use of fecal MPO was selected as a complementary objective marker of neutrophil-associated intestinal inflammation because it is highly responsive to dietary modulation in both experimental and human intervention studies^10, 15-17^. Moreover, MPO may detect short-term inflammatory changes that are not readily captured by biomarkers typically monitored over longer clinical intervals, such as FC.

Secondary outcomes focused on changes to fecal microbiota, as well as the proportion of patients who intended to continue the study diet when food was no longer provided without cost, and the reasons for discontinuation of either diet (compared to resuming usual food habits). Additional dichotomous outcomes included body weight, HBI, FC (<50ug/g normal, >250ug/g inflammation) and hsCRP (<1.0gm/L normal, >5mg/L inflammation). The HBI includes general well-being, abdominal pain, number of liquid stools per day, presence of an abdominal mass, and complications, with scores of ≤4 indicating remission, and a score >8 indicating moderate disease. To complement CDAI assessment, daily patient-reported symptoms were captured using the short CDAI (sCDAI) via REDCap survey starting from 7 days before diet initiation through completion of the study intervention, including diarrhea, abdominal pain, and general well-being. A dietitian-administered 24-hour dietary recall was performed at enrollment and at time of baseline stool collection. A food frequency questionnaire was conducted at enrollment (Diet History Questionnaire II, National Cancer Institute)^18^.

### Diet Adherence

Adherence was measured via daily REDCap survey asking “If you consumed any foods yesterday that were not provided to you by this study, please describe/list them here”. At study completion, participants also provided a log of the provided study-provided foods consumed and not consumed and completed a 24-hour dietary recall in-person with a trained dietitian.

### Adverse Events

Adverse events were assessed at the end of study visit (day 7 of diet). Serious adverse events included any adverse event that was fatal, life-threatening, requiring prolonged hospital stay, congenital anomaly or birth defect or other medically significant event as deemed such by the investigator.

### Protocol Deviations and Monitoring

The **Supplementary Materials** describe the protocol deviations, modifications, and monitoring and data quality.

### Ethical Considerations

The Institutional Review Board of University Hospitals Cleveland Medical Center gave ethical approval for this work (STUDY20190080). All patients provided written informed consent before study entry and re-consent was obtained if participating for a second time. All authors had access to the study data and reviewed and approved the final manuscript.

The study protocol, including design, variables, treatment conditions, and statistical analysis plan were registered prior to conducting the research on ClinicalTrials.gov (NCT04065048) on August 20, 2019. Initial registration (August 20, 2019) on ClinicalTrials.gov, based on a pilot feasibility grant, specified a primary outcome of change in CDAI from baseline (d0) to post-intervention (d7); following subsequent NIH funding, the registry was updated on December 16, 2020 to include quantification of fecal MPO. The study followed CONSORT guidelines (see **Supplementary Materials**; CONSORT checklist).

### Data Sharing Statement

Deidentified individual participant data are available indefinitely at figshare doi:10.6084/m9.figshare.30075568. The study protocol is available on ClinicalTrials.gov (NCT04065048).

### Fecal Microbiota Transplantation Experiments in Mice

To understand the role of microbiome changes on inflammation, we conducted a series of fecal microbiota experiments. Groups of six 14-week-old sex-matched (littermate controls) inbred germ-free (GF) SAMP1/YitFC (SAMP) mice (Cleveland Digestive Diseases Research Center, Mouse Models Core) transplanted with either ‘pre’ (d0) or ‘post’ (d7) SoyP or AnimalP diet-altered human feces were used to quantify the inflammatory (functional) potential diet-altered human gut microbiota. Details on the human gut microbiota transplanted GF SAMP (hGM-SAMP) model is previously described^19^. Two weeks post colonization, colitis was induced with 3% DSS (TdB Consultancy AB) offered ad libitum for 7 days and mice resumed with water for 2 days, then were killed. The impact of FMT was assessed using five parameters associated with colonic inflammation (MPO, FITC, colon length, and histology). Animals were maintained on a standard rodent chow following weaning. Experimental methods are described in the **Supplementary Materials**. All procedures were approved by the Institutional Animal Care and Use Committee and the Institutional Review Board at CWRU.

### Human Fecal Microbiome Analysis

Fecal DNA was extracted using the ZymoBIOMICS Mini Prep Kit, and DNA libraries were generated with the Illumina Nextera XT library preparation kit, following a modified protocol. Library concentrations were measured using the Qubit fluorometer (ThermoFisher). Sequencing was performed on an Illumina HiSeq platform with a read length of 2 × 150 bp. The unassembled sequencing reads were processed and analyzed through the CosmosID bioinformatics platform (CosmosID Inc.), as previously described^19-23^.

### Statistical Methods and Power

Descriptive data are reported as mean and SD or as counts and percentages. Formal statistical comparisons of descriptive variables and within group analysis comparing baseline (d0) and post-intervention (d7) values were performed using the Wilcoxon signed-rank test for continuous variables and the chi squared or Fisher’s exact test for categorical variables. The primary analysis compared the absolute effect before/after (Δd7-d0) treatment effect using Mann-Whitney U t-test or comparable non-parametric test for ANOVA comparison across groups. To control for outcome multiplicity (multiple hypothesis testing) we used a highly conservative family wise error rate (FWER) multiple hypothesis correction applying Bonferroni adjustment to adjust the p-values for the two primary outcomes.

Risk ratios and corresponding 95% confidence intervals with P-values (two-sided Fisher exact) are also reported. We included both male and female participants. Although blinding of study participants and evaluators to treatment assignment was not feasible, all sample analyses and statistical evaluations were conducted in a blinded fashion. Analysis was performed using GraphPad Prism 10 and STATA software (v15.1).

To account for the potential confounding effect of sex, we conducted multivariable regression analyses (linear regression and generalized linear models) with MPO as the dependent variable and sex, diet, disease status, and other covariates as independent variables. Potential interaction effects were assessed by evaluating whether inclusion of covariates meaningfully altered the association between diet and inflammatory outcomes. Variables that did not materially influence model estimates were excluded from the final model. The effect of sex was reported using p-values and 95% confidence intervals. In addition, we performed a dichotomous analysis to evaluate the effect of diet on changes in MPO, classifying responses as beneficial or adverse. Risk ratios and corresponding 95% confidence intervals are reported. As an exploratory post hoc analysis, a rank-based composite outcome (CDAI-MPO) was constructed to capture the combined effects of diet on clinical disease activity and intestinal inflammation using changes in CDAI and fecal MPO from baseline (d0) to post-intervention (d7). For each participant, CDAI and MPO change values were categorized into five ordered levels (1-5) reflecting the magnitude and direction of change. CDAI categories were defined as: 1; Largest reduction (≤ -88), 2; Moderate reduction (-87 to -34), 3; Mild reduction / near neutral (-33 to -8), 4; Neutral / slight increase (-7 to 5), and 5; Increase (≥ 9). MPO categories were defined as: 1; Largest reduction (≤ -1.535), 2; Moderate reduction (-1.534 to - 0.877), 3; Mild reduction / near neutral (-0.876 to -0.238), 4; Neutral / slight increase (-0.237 to 0.381), and 5; Increase (≥ 0.639). Lower category values indicate greater improvement. A composite score was calculated as the sum of CDAI and MPO category ranks (range 2–10), with lower scores representing more favorable overall responses. Between-group differences in the composite score were evaluated using the Mann-Whitney U test, and effect size was estimated using the Hodges-Lehmann method with exact 95% CIs.

Sample size estimations were originally based on the mean±SD computed from the mouse fecal MPO data in human gut microbiota transplanted GF SAMP groups treated with the soy/pea or chow diet for the SAMP mouse model of CD-ileitis^24^. Power tables indicated that 16 subjects per diet would be sufficient to reach a power of 97% provided that the magnitude of the effect of the diet on human fecal MPO is similar to that seen in mice (relative reduction by 50%), which in the mouse model was representative of about 13.4µg reduction over 7 days compared to the reduction of -0.9µg in animals receiving the chow diet.

## RESULTS

### Participants

In total, 572 CD individuals were contacted via recruitment letter. Of these, 81 (14%) expressed interest, of whom 47 (48%) were deemed ineligible. A total of 79 HC participants expressed interest in the study, of whom 47 (59%) were deemed ineligible. The patient flow chart is shown in **Figure 1**, summarizing recruitment, diet allocation, and early discontinuation. Six female participants withdrew from the trial, 5 of whom prior to starting the diet, and 2 due to adverse events.

**Figure 1.**
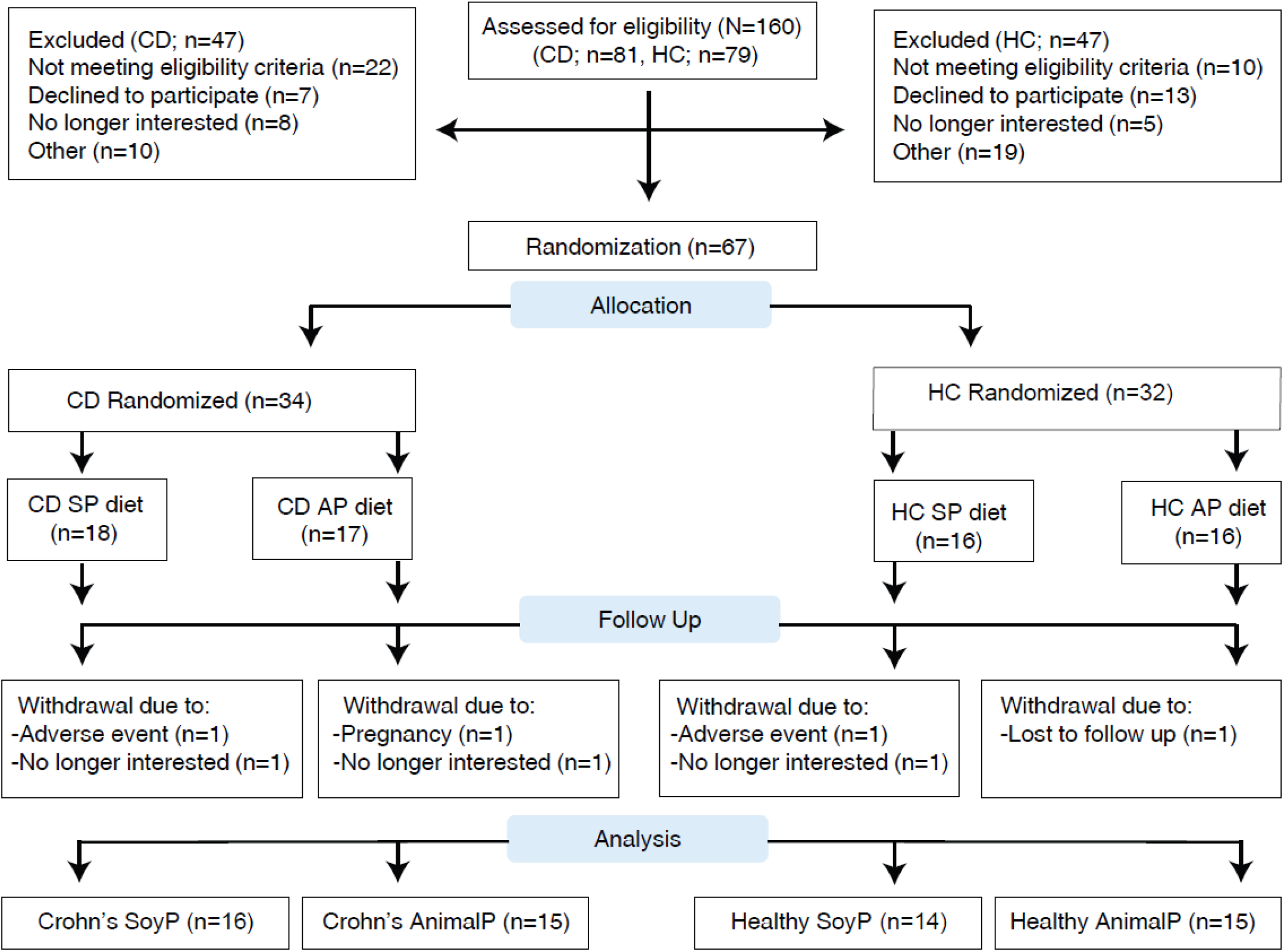
Study enrollment, allocation of diets, and follow-up.

Baseline characteristics of all study participants are shown in **Table 1**. Covariates were generally well balanced between CD and HC and between treatment groups, except for age, wherein CD participants were significantly older. Overall, 68% (41) of the participants were women, and majority of participants were White, followed by African American and Asian. The median body mass index at screening was 28.2 kg/m^2^ (IQR 24.97, 31.99 kg/m^2^). Among the CD participants at enrollment, no difference in number of years since CD diagnosis, disease characteristics or medication use was identified between the diet groups (**Table 2**). Although there was a small reduction in weight loss across all groups, no differences were observed in the mean percentage change in weight (lbs) across all diet study groups (One-way ANOVA P=0.71, CD-SoyP -0.59±1.6; HC-SoyP -1.37±1.5; CD-AnimalP, -1.18±2.4, and HC-AnimalP, -0.99±1.5; pairwise difference of means across groups was <0.7495%).

**Table 1.** Baseline Characteristics of Study Participants.

| Characteristic | Crohn's, N=31 | Healthy, N=29 | P-value* |
| --- | --- | --- | --- |
| Age at randomization, y | 47.45 ± 12.21 | 35.6 ± 13.3 | 0.0007 |
| Female sex, n (%) | 18 (58) | 23 (79.3) | 0.099 |
| Race, n (%) |  |  |  |
| Asian | 0 (0) | 5 (17.2) | 0.021 |
| Black | 3 (9.7) | 4 (13.9) | 0.702 |
| White | 28 (90.3) | 20 (68.9) | 0.054 |
| Ethnicity, n (%) |  |  |  |
| non-Hispanic | 31 (100) | 28 (96.5) | 0.483 |
| BMI at baseline, kg/m<sup>2</sup> | 29.53 ± 6.54 | 28.68 ± 5.68 | 0.591 |

\*Fischer's exact or Chi-square statistics p. Welch's t-test used to compare means and SD.

**Table 2.**
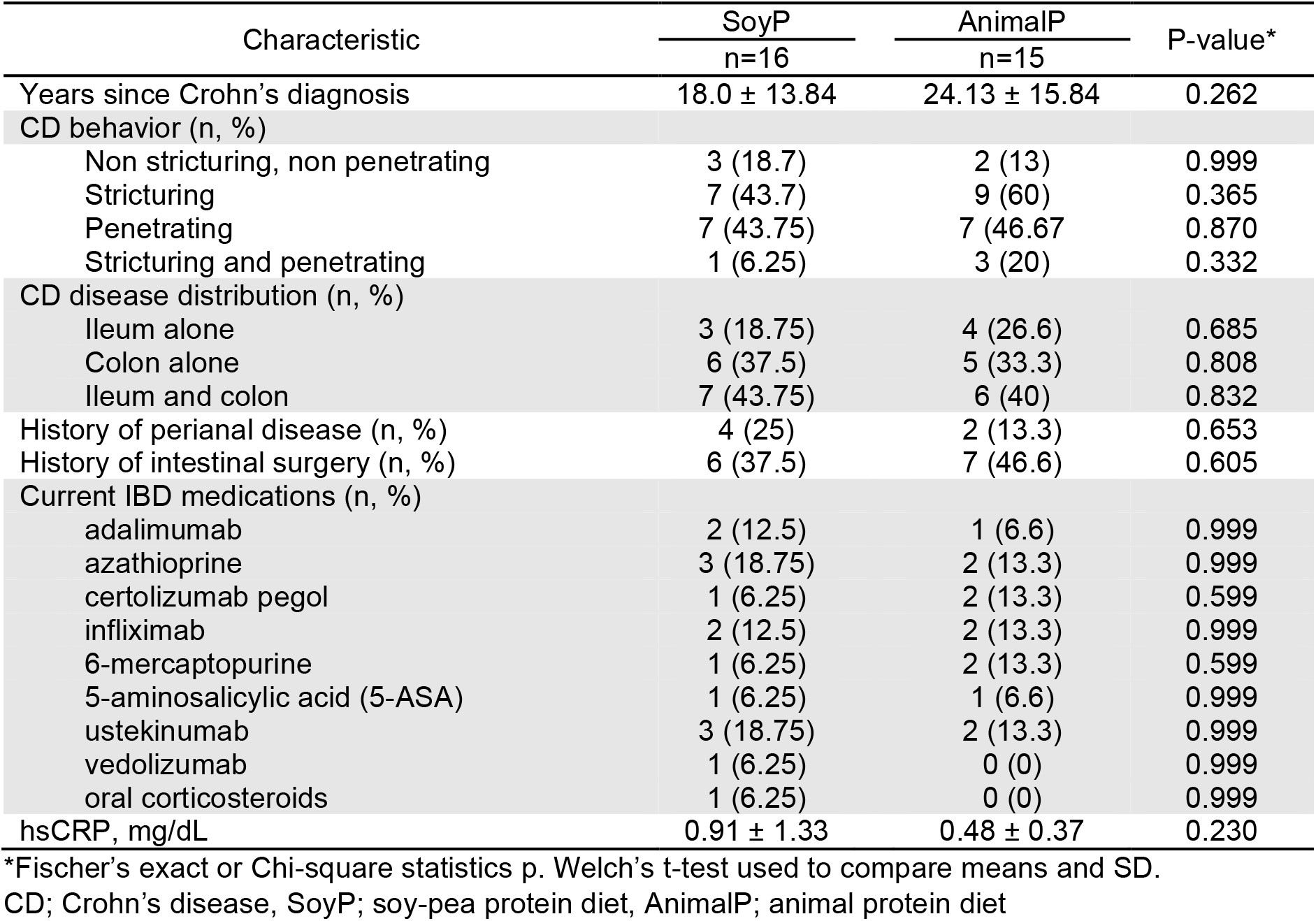
Baseline Characteristics of Crohn’s Disease Participants.

| Characteristic | SoyP<br>n=16 | AnimalP<br>n=15 | P-value* |
| --- | --- | --- | --- |
| Years since Crohn's diagnosis | 18.0 ± 13.84 | 24.13 ± 15.84 | 0.262 |
| CD behavior (n, %) |  |  |  |
| Non stricturing, non penetrating | 3 (18.7) | 2 (13) | 0.999 |
| Stricturing | 7 (43.7) | 9 (60) | 0.365 |
| Penetrating | 7 (43.75) | 7 (46.67) | 0.870 |
| Stricturing and penetrating | 1 (6.25) | 3 (20) | 0.332 |
| CD disease distribution (n, %) |  |  |  |
| Ileum alone | 3 (18.75) | 4 (26.6) | 0.685 |
| Colon alone | 6 (37.5) | 5 (33.3) | 0.808 |
| Ileum and colon | 7 (43.75) | 6 (40) | 0.832 |
| History of perianal disease (n, %) | 4 (25) | 2 (13.3) | 0.653 |
| History of intestinal surgery (n, %) | 6 (37.5) | 7 (46.6) | 0.605 |
| Current IBD medications (n, %) |  |  |  |
| adalimumab | 2 (12.5) | 1 (6.6) | 0.999 |
| azathioprine | 3 (18.75) | 2 (13.3) | 0.999 |
| certolizumab pegol | 1 (6.25) | 2 (13.3) | 0.599 |
| infliximab | 2 (12.5) | 2 (13.3) | 0.999 |
| 6-mercaptopurine | 1 (6.25) | 2 (13.3) | 0.599 |
| 5-aminosalicylic acid (5-ASA) | 1 (6.25) | 1 (6.6) | 0.999 |
| ustekinumab | 3 (18.75) | 2 (13.3) | 0.999 |
| vedolizumab | 1 (6.25) | 0 (0) | 0.999 |
| oral corticosteroids | 1 (6.25) | 0 (0) | 0.999 |
| hsCRP, mg/dL | 0.91 ± 1.33 | 0.48 ± 0.37 | 0.230 |

\*Fischer's exact or Chi-square statistics p. Welch's t-test used to compare means and SD.
CD; Crohn's disease, SoyP; soy-pea protein diet, AnimalP; animal protein diet

### Primary and Secondary Outcomes

#### Effect on Patient-Reported Symptoms by CDAI and HBI scores

##### CDAI and HBI score

The final analysis included 60 participants (CD n=31, HC n=29) (**Figure 2A**). Change in CD symptoms (CDAI, HBI, sCDAI) from baseline to day 7 for the study diet groups is shown in **Table 3**. After 7 days of offering the diets to participants, significantly more of the CD-SoyP diet participants experienced a reduction in CDAI and HBI score, an effect not seen among the CD-AnimalP diet group (**Figure 2B**). The SoyP diet did not result in a significant between-group difference in the change (Δd7-d0) in CDAI score compared with the AnimalP diet group (**Figure 2C, Table 3**), although risk-ratio analysis demonstrated a directional trend favoring SoyP (RR = 1.52, 95% CI: 0.89-2.58, P = 0.135). No CD participants experienced worsening of CDAI score. While the between-group difference in CDAI did not reach statistical significance, the change (Δd7-d0) in HBI score was significantly greater in the SoyP diet group compared with the AnimalP group (**Figure 2C, Table 3**). Risk-ratio analysis showed that CD participants assigned to the SoyP diet were 4.68 times more likely to experience a reduction in HBI score compared with those assigned the AnimalP diet (RR = 4.68, 95% CI: 1.22-17.98, P = 0.009).

**Table 3.**
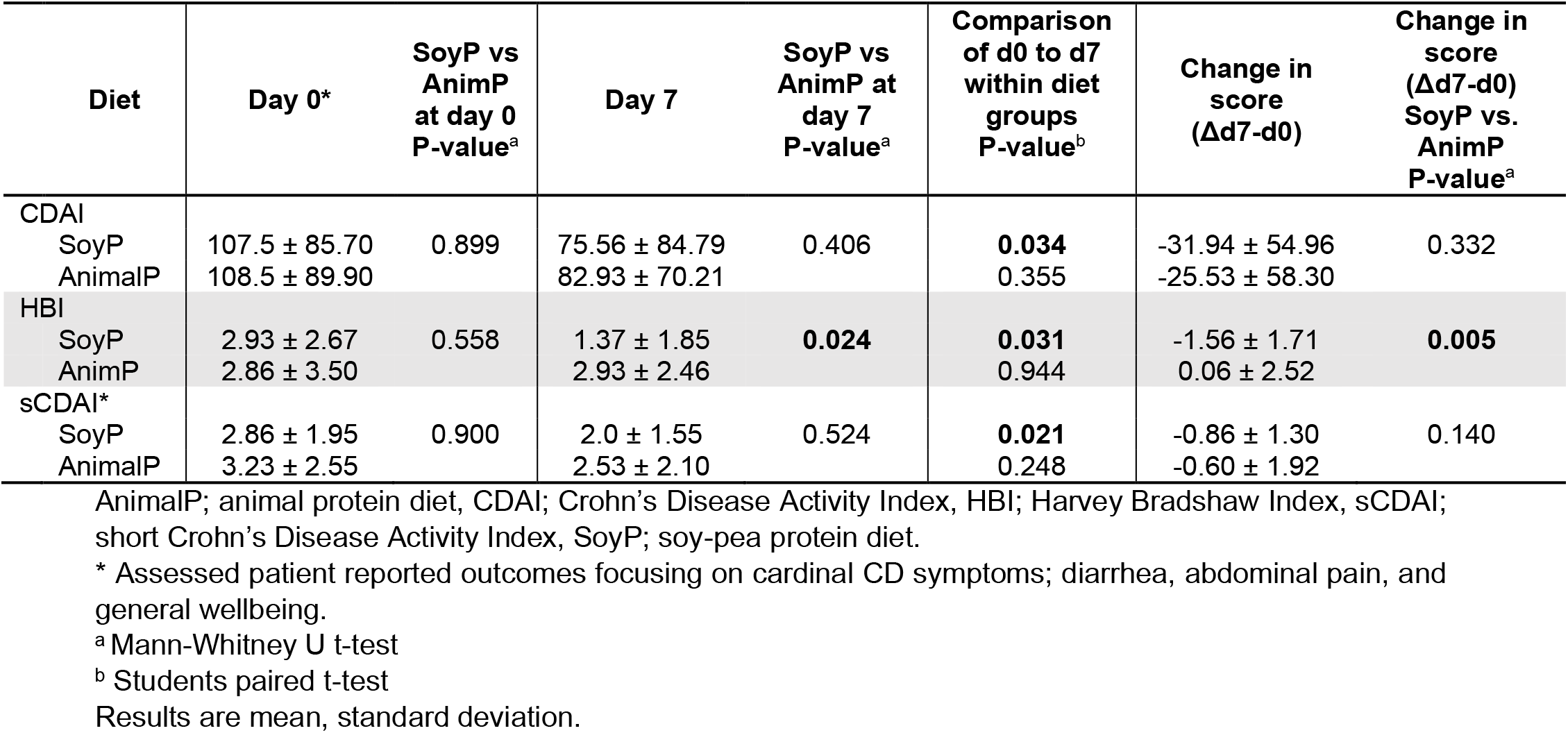
Change in Symptoms from Day 0 to day 7 on the Study Diet in Crohn’s Disease Participants.

**Figure 2.**
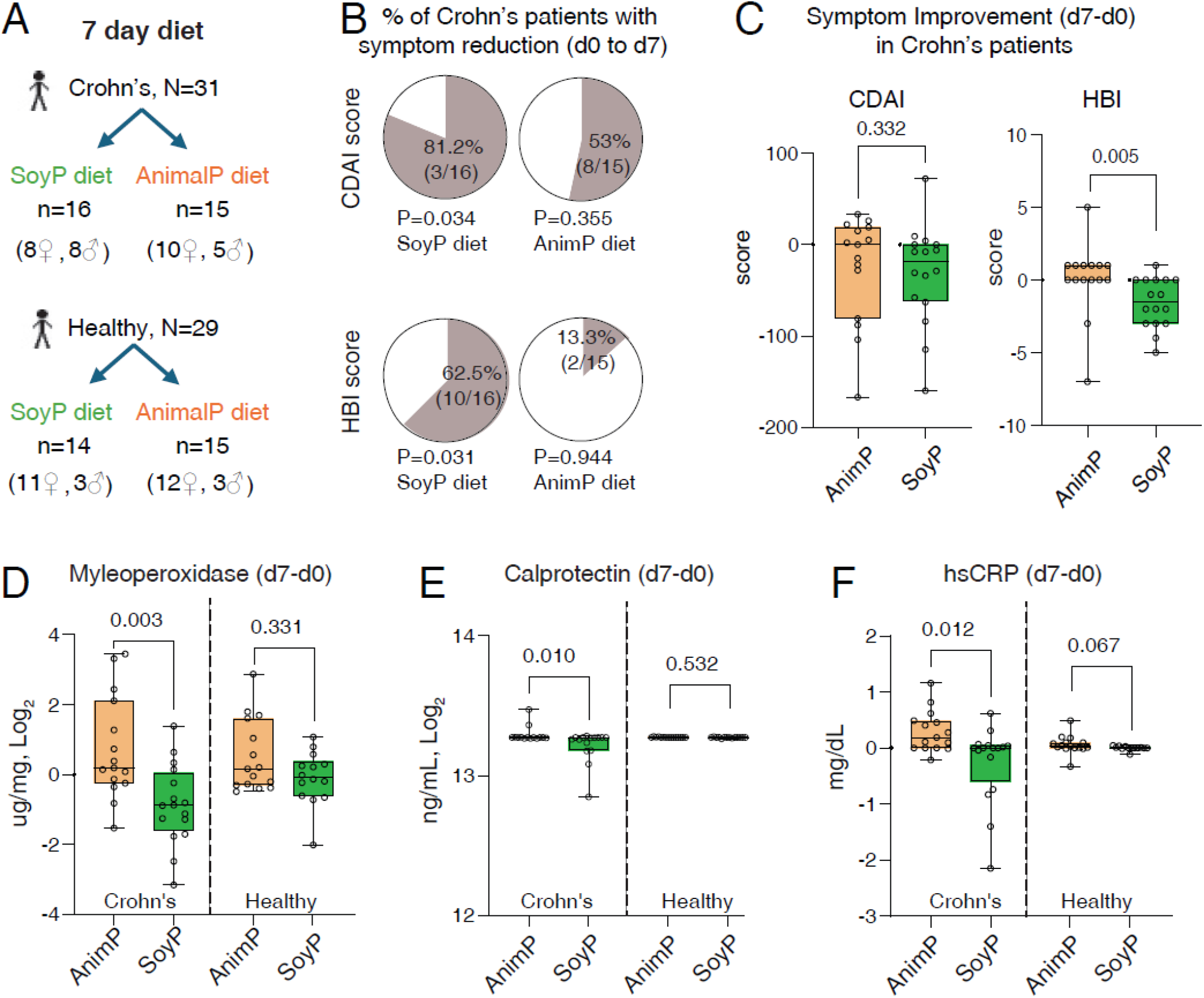
Soy-pea protein diet reduces inflammatory markers in patients with Crohn’s disease. Change in inflammatory markers (Δd7-d0) for CD vs HC participants. **A)** Participant diet allocation by sex. **B)** Pie charts illustrate proportion of CD participants who experienced reduction (shaded area) in CDAI and HBI score. P-value is paired t-test comparison of mean±SD for d0 and d7 within diet groups. **C)** Change (d7-d0) in score/value and representative Mann Whitney statistics p-value for; Crohn’s disease activity index (CDAI) and Harvey Bradshaw index (HBI), **D)** fecal myeloperoxidase, **E)** fecal calprotectin, and **F)** C-reactive protein.

##### sCDAI score

A significant reduction in sCDAI score was observed from d0 to d7 among 56.2% (9/16) of CD participants on the SoyP-diet (Paired t-test = 0.021), compared to 4/15 (26.6%) on the AnimalP-diet (Paired t-test P = 0.248) (**Table 3**).

#### Markers of Inflammation

##### Fecal Myleoperoxidase (MPO)

Analysis of biochemical inflammation in the feces (MPO, FC) and serum (hsCRP) of participants at the start of the study diet (d0) and day 7 of the diet is shown in **Table 4**. Despite randomization, there were significant differences in the baseline fecal MPO values between the CD diet groups.

**Table 4.** Response of markers of inflammation to a diet rich in soy and pea compared to Animal protein diet from Day 0 to Day 7 in Crohn’s Disease and Healthy Control Participants.

| Group | Diet | Day 0 | P-value<sup>a</sup> | Day 7 | Participants with reduction (d0 to d7), n (%) | Within group by diet (d0 to d7) P-value<sup>c</sup> | Change (Δd7-d0) | Within group by diet (Δd7-d0), P-value<sup>d</sup> | Between group by diet (Δd7-d0), P-value<sup>e</sup> |
| --- | --- | --- | --- | --- | --- | --- | --- | --- | --- |
| Myeloperoxidase (ug/mg, Log<sub>2</sub>)<sup>b</sup> |  |  |  |  |  |  |  |  |  |
| Crohn's | SoyP | 4.36 ± 2.00 | **0.006<sup>b</sup>** | 3.50 ± 2.05 | 12/16 (75) | **0.009** | -0.89 ± 1.15 | **0.003** | **0.027** |
|  | AnimalP | 2.65 ± 1.55 |  | 3.38 ± 1.68 | 5/15 (33.3) | 0.076 | 0.73 ± 1.48 |  |  |
| Healthy | SoyP | 2.24 ± 1.14 |  | 2.14 ± 1.09 | 8/14 (57.1) | 0.635 | -0.09 ± 0.76 | 0.331 | 0.653 |
|  | AnimalP | 2.46 ± 0.83 |  | 2.98 ± 1.12 | 7/15 (46.6) | 0.072 | 0.52 ± 1.03 |  |  |
| Fecal calprotectin (ng/mL) |  |  |  |  |  |  |  |  |  |
| Crohn's | SoyP | 620.9 ± 1097 | 0.101 | 257.2 ± 434.0 | 13/16 (81.2) | 0.063 | -363.7 ± 697.4 | **0.011** | **0.010** |
|  | AnimalP | 205.7 ± 312.7 |  | 343.8 ± 724.4 | 6/14 (42.8) | 0.237 | 138.0 ± 417.1 |  |  |
| Healthy | SoyP | 95.54 ± 27.16 |  | 90.42 ± 10.25 | 7/14 (50) | 0.420 | -5.12 ± 23.01 | 0.532 | 0.533 |
|  | AnimalP | 88.51 ± 6.85 |  | 93.91 ± 20.43 | 5/15 (33.3) | 0.176 | 5.39 ± 14.68 |  |  |
| hs C-reactive protein (mg/dL) |  |  |  |  |  |  |  |  |  |
| Crohn's | SoyP | 0.91 ± 1.33 | 0.167 | 0.65 ± 0.82 | 6/16 (37.5) | 0.150 | -0.26 ± 0.69 | **0.012** | 0.667 |
|  | AnimalP | 0.48 ± 0.37 |  | 0.77 ± 0.64 | 2/15 (13.3) | **0.008** | 0.29 ± 0.36 |  |  |
| Healthy | SoyP | 0.25 ± 0.24 |  | 0.25 ± 0.23 | 1/14 (7.1) | 0.887 | 0.001 ± 0.03 | 0.067 | 0.113 |
|  | AnimalP | 0.36 ± 0.41 |  | 0.42 ± 0.51 | 3/15 (20) | 0.179 | 0.06 ± 0.16 |  |  |

<sup>a</sup> Kruskal-Wallis, comparison of d0 values between CD and HC diet groups.
<sup>b</sup> Day 0 fecal MPO was higher for the CD-SoyP than CD-AnimalP diet group (P=0.012). No difference identified between HC diet groups (P= 0.651).
<sup>c</sup> Students paired t-test, comparison of d0 to d7 within groups by diet.
<sup>d</sup> Mann-Whitney U test (Δd7-d0); CD-SoyP vs CD-AnimalP, HC-SoyP vs HC-AnimalP.
<sup>e</sup> Mann-Whitney U test (Δd7-d0); CD-SoyP vs HC-SoyP, CD-AnimalP vs HC-AnimalP.
Results are mean, standard deviation.

Overall, paired analysis of day 0 and day 7 showed a significant within-group reduction in fecal MPO for the CD-SoyP participants (75%, 12/16, P=0.009), which was not seen among the other diet groups (**Table 4**). Between-group comparisons demonstrated a significant difference in the change in MPO (Δd7-d0) between the CD-SoyP and CD-AnimalP groups (P = 0.003), as well as between the CD-SoyP and HC-SoyP groups (P = 0.027) (**Figure 2D, Table 4**).

To account for baseline imbalance and potential regression-to-the-mean effects, an ANCOVA adjusting for baseline MPO values was performed, and the difference between the CD-SoyP and CD-AnimalP groups remained significant (ANCOVA: β = -1.21, 95% CI: -2.27 to -0.16, P = 0.025). The effect of diet on fecal MPO remained statistically significant after Bonferroni adjustment for the two prespecified primary outcomes (FWER adjusted P = 0.006), while the CDAI remained non-significant (FWER adjusted P = 0.664).

Risk ratio analysis showed that CD patients assigned the SoyP diet were more likely than the CD-AnimalP diet group to experience a reduction in fecal MPO (RR=2.3, 95% CI: 1.04-4.85, P=0.032). The effect of diet on fecal MPO in CD and HC groups was estimated as RR = 2.10 (95% CI: 0.98-4.48, P=0.063) for the SoyP diet and RR = 0.71 (95% CI: 0.29-1.75, P=0.710) for the AnimalP diet.

##### Fecal Calprotectin (FC)

At the start of the study diet no difference in FC concentrations (ng/mL) between CD and HC diet groups was identified. Additionally, there was no difference in the proportion of CD participants who had a FC level >120ng/ml or >600ng/ml by study diet (Fishers Exact P=0.38, P>0.99, respectively), or of HC participants by study diet (P>0.99 for both).

When examining the effect of diet on FC, risk-ratio analysis showed CD-SoyP vs CD-AnimalP (RR = 1.89, 95% CI: 0.99-3.62, P=0.056), CD-SoyP vs HC-SoyP (RR = 1.60, 95% CI: 0.91-2.88, P=0.077), and for CD-AnimalP vs HC-AnimalP groups (RR = 1.60, 95% CI: 0.67-3.77, P=0.462). Overall, 81.2% (13/16) of CD participants on the SoyP diet had a reduction in FC from day 0 to day 7 (**Table 4**). Although dichotomous risk-ratio analysis did not reach conventional statistical significance, continuous change analysis demonstrated significant between-group differences in FC (Δd7-d0) between the CD-SoyP and CD-AnimalP groups (**Figure 2E, Table 4**).

##### Serum high-sensitivity C-Reactive Protein (hsCRP)

Although the hsCRP values for the CD and HC groups were within a threshold of 5mg/dL, as shown in Table 2, the average hsCRP in CD participants was 2.3 times higher than that in HC participants.

When examining the effect of diet on systemic inflammation (hsCRP), risk-ratio analysis showed CD-SoyP vs CD-AnimalP (RR = 2.8, 95% CI: 0.66-11.83, P = 0.22), CD-SoyP vs HC-SoyP (RR = 5.25, 95% CI: 0.71-38.47, P = 0.086), and CD-AnimalP vs HC-AnimalP (RR = 0.66, 95% CI: 0.13-3.43, P = 1.00). Notably, among CD participants, 37.5% (6/16) in the SoyP group had a reduction in hsCRP from day 0 to day 7, whereas 86.6% (13/15) in the AnimalP group experienced an increase (**Table 4**). This opposing directional response contributed to a significant difference in change (Δd7-d0) between the CD-SoyP and CD-AnimalP groups (Mann-Whitney P = 0.012; **Figure 2F, Table 4**).

##### Composite CDAI-MPO Analysis

As an exploratory supportive post-hoc analysis, MPO and CDAI change values were rescaled and categorized into quintiles reflecting the magnitude and direction of effect (1 - ‘largest reduction’ to 5 - ‘increase’), and a composite CDAI-MPO score was derived. The composite CDAI-MPO score was significantly lower in the SoyP group compared with the AnimalP group (median 5 [IQR 4-6] vs 8 [IQR 7-9], Mann-Whitney U = 56.5, P = 0.012). Participants in the SoyP group were more likely to fall within categories 1-2 (‘large’ to ‘moderate’ reduction) compared with the AnimalP group (RR = 4.69, 95% CI: 1.22-4.68, P = 0.009), whereas assignment to category 5 (‘increase’) was less frequent (RR = 0.31, 95% CI: 0.07-1.31, P = 0.113).The Hodges-Lehmann estimate indicated a 3-point lower composite score in the SoyP group (-3.0; 95% CI: -3 to -1).

##### Effect of diet is independent of sex effect

To assess to what extent the effect of diet on inflammation was influenced by sex as a variable, we conducted multivariable regression analysis to quantify the interaction of inflammatory markers, diet and sex. Overall, when data from both CD and HC were included in the model, sex had no significant interaction effect with MPO, HBI, hsCRP and FC (i.e., p values remained unchanged in all models that excluded the variable sex). On regression analysis, the SoyP-diet showed significant effects reducing fecal MPO (P=0.0001), HBI scores (P=0.034) and hsCRP (P<0.014), with no interaction for sex (P>0.71). Analysis of these significant variables (MPO, HBI, hsCRP), conducted for each CD and HC groups separately, showed that the effect observed on inflammation markers was due to the diet, and not due to sex, or other demographic variables. For instance, when regression analysis was restricted to CD individuals, the SoyP-diet was associated with a greater reduction in fecal MPO (P<0.0001; coefficient = -1.752, 95%CI: -2.272 to -0.778) and hsCRP (P=0.006; coefficient= -0.55, 95%CI: -0.95 to -0.162) with no effect of sex (P=0.794 and P>0.713, respectively). Removal of sex from the model did not alter these estimated coefficients. Among HC participants, the multivariable effect of the SoyP-diet on fecal MPO slightly changed from P=0.073 (coefficient= -0.615, 95%CI: -1.287 to 0.056; with sex P=0.415) to P=0.069 with nearly identical coefficients (-0.62, 95%CI: -1.28 to 0.047), confirming that the effect of the diet was not influenced by sex. Overall, the direction and magnitude of MPO reduction by the SoyP-diet was substantially greater in CD. Similarly, the influence of sex on the effect of diet on other fecal markers, such as FC, was determined to be nonsignificant (P>0.453).

Overall, these findings indicate that the observed reductions in inflammatory markers were driven by dietary intervention rather than sex or other demographic variables, with effects most pronounced among participants with CD.

##### CDAI-stratified analysis reveals differential dietary response by disease activity

To explore whether baseline disease activity influenced response to the diet, participants with CD were stratified by CDAI score, with a CDAI <150 indicating remission. Regression analysis based on patients with CDAI <150 at study commencement vs those with CDAI>150, revealed that the statistical association between SoyP diet consumption and fecal MPO activity (ug/mg, Log^2^) remained significant in patients with CDAI <150 (P<0.0001; coefficient= -2.58, 95%CI: -3.56 to -1.60). Of interest, this association became non-significant in patients with CDAI greater than 150 (P=0.799). The lack of significance in this group appeared to result from heterogeneity in response, with some individuals showing a positive MPO response to SoyP and others showing no (neutral) response. Healthy controls on the SoyP diet exhibited a modest but non-significant trend toward MPO reduction (P=0.069; coefficient= -0.62, 95%CI: -1.28 to 0.04), consistent with lower baseline inflammatory burden.

### Participation for a second time

Of the 60 total participants, 10 CD and 5 HC individuals participated in the study for a second time and received the other diet that they did not receive the first time. Paired analysis of the change in fecal MPO (Δd7-d0) among CD and HC participants who completed both diet arms are shown in **Figure 3**. Overall, CD participants had a greater improvement in fecal MPO with the SoyP-diet compared to when the same participants were assigned AnimalP-diet, an effect not seen among HC participants (**Figure 3A-D**). No significant difference in hsCRP response was observed among CD or HC participants who completed both diet arms (**Figure 3E-F**). When looking at disease symptoms scores, the SoyP diet was found to have a positive effect on symptoms, with 80% (8/10) of the CD-SoyP participants experiencing a reduction in HBI score compared to 10% (1/10) when assigned the AnimalP-diet (**Figure 3G-H**).

**Figure 3.**
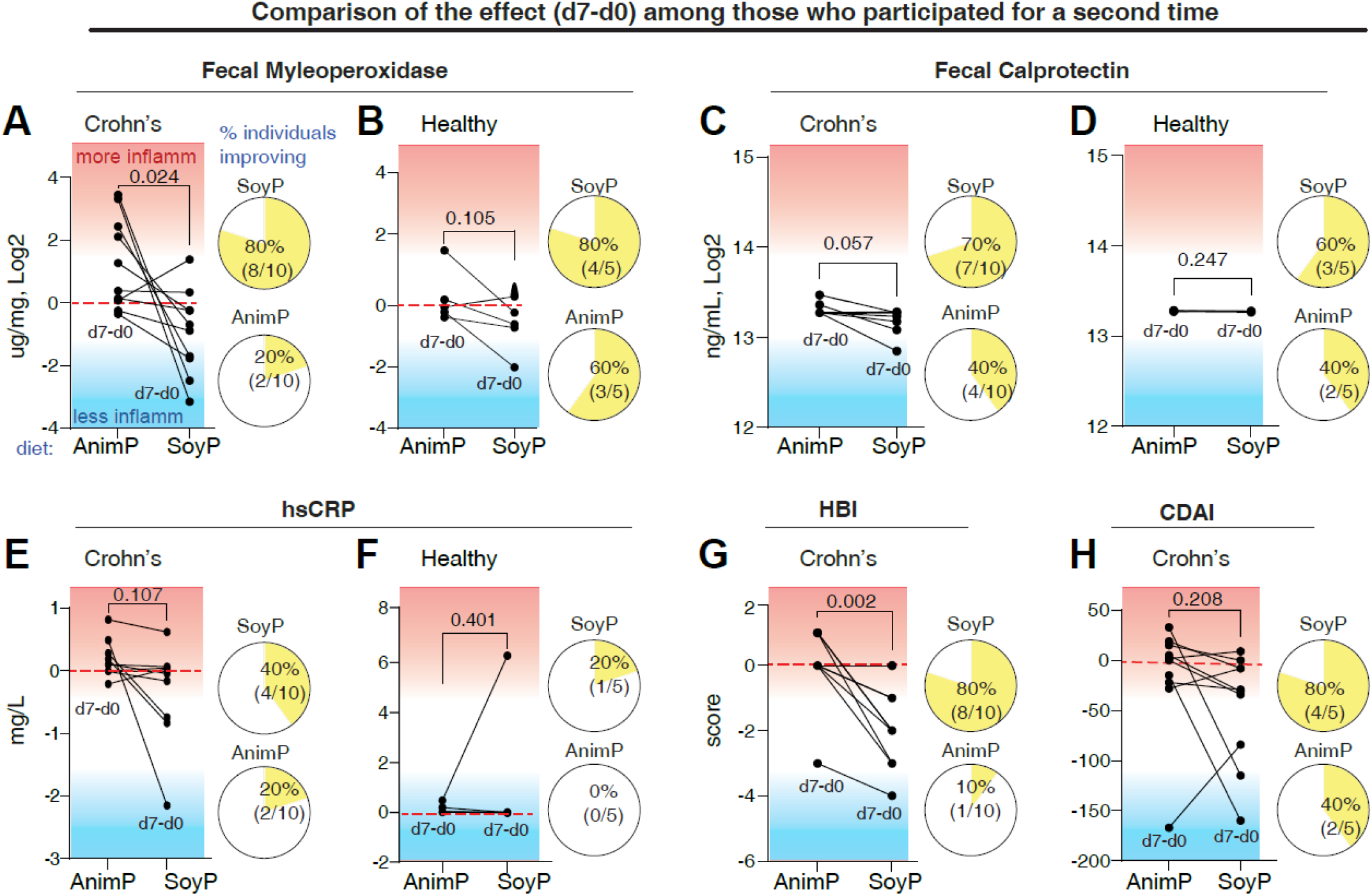
Participation in the study for a second time showed greater improvement in inflammatory parameters in patients on the SoyP-diet. Paired analysis of CD (n=10) and HC participants (n=5) who completed the diet for a second time. Each graph compares the change (d7-d0) in score/value for AnimalP-diet (left side) and SoyP-diet (right side). Pie charts illustrate the proportion of individuals who experienced a reduction in outcome measure. A-B) Fecal myeloperoxidase, C-D) Fecal calprotectin, and E-F) hsCRP for CD and HC, respectively. G-H) Harvey Bradshaw index; HBI and Crohn’s disease activity index; CDAI.

#### Most patients opted to continue consuming Soy-Pea after the end of study

Six months following the completion of the study, participants were sent a questionnaire asking who continued the study diets when food is no longer provided without cost. Survey response rates were higher among participants completing the SoyP-diet [CD; 75%(12/16), HC; 50%(7/14)] compared with those on the AnimalP-diet [CD; 53% (8/15), HC; 40% (6/15)]. Overall, across all participants assigned to the SoyP diet, 63.1% (12/19) reported that they would continue to include soy/pea protein in their diet, compared with 42.8% (6/14) of those who completed the AnimalP-diet (Chi-square, P=0.304). Analysis based on diet groups showed more participants reported that they would continue to include soy/pea protein in their diet (66%, 8/12 of CD, vs. 57%, 4/7 of HC, Fisher’s Exact, P=1.0) than participants reporting that they would continue an animal protein-based diet (62.5%, 5/8 of CD, vs. 16.7%, 1/6 of HC, Fisher’s Exact, P=0.137).

### Adherence and Macronutrient Intake

Overall, only one participant on the SoyP-diet reported a single occurrence of consuming a non-compliant food item (**Supplementary Materials**). Foods consumed that were not provided by the study are shown in **Supplementary Tables 1-2**. Mean total macronutrient intake (energy, fat, carbohydrate, and protein) did not differ between or within study groups, confirming that the diets were well matched. Fiber intake was higher among male CD-SoyP compared to CD-AnimalP participants (34.6 ± 8.1 vs 22.2 ± 6.7, p = 0.017). As expected based on dietary composition, intake of soy-derived isoflavones, including daidzein, genistein, and glycitein, was higher in all SoyP vs AnimalP diet groups (all P < 0.001). Other micronutrient differences reflected the underlying nutrient profiles of plant versus animal protein sources, including higher folate, iron, and magnesium in SoyP groups, and higher vitamin B12, methionine, and selenium in AnimalP groups. Comparison of total macronutrient and micronutrient intake within and between groups are shown in **Supplementary Tables 3-14**.

### Adverse events

The diets were well-tolerated over the 7 days. Most of the adverse events reported were gastrointestinal related (e.g., flatulence, bloating) and resolved within the first 1-2 days. No serious adverse events were reported. See **Supplementary Materials** for details.

### Diet-Associated Changes in Fecal Microbial Profiles and Functional Pathways

We next conducted metagenome sequencing of pre and post fecal samples to determine how the diets affect the gut microbiome. Although the relative abundance (Shannon diversity) across CD participants was lower than HC before the study initiated (P< 0.05), alpha diversity analysis showed no differences due to either diet among CD or HC participants after 7 days (**Figure 4A**). Overall, at the phylum level, gut microbial composition across all participants irrespective of diet or sampling time (days 0 or 7) was enriched for *Proteobacteria* and *Firmicutes* (mainly *Clostridia*), followed by *Bacteroidetes* (mainly *Bacteroidia*) (**Figure 4B**). At the family level, compared to HC, CD participants had less *Ruminococcaceae, Veillonellaceae, Lachnospiraceae*, and *Prevotellaceae* and more *Bacteroidaceae*. **Figure 4C** illustrates that family-level differences were similar for various groups of participants that were recruited as cohorts at different times (i.e., cohorts of 14-16; N=60 total).

**Figure 4.**
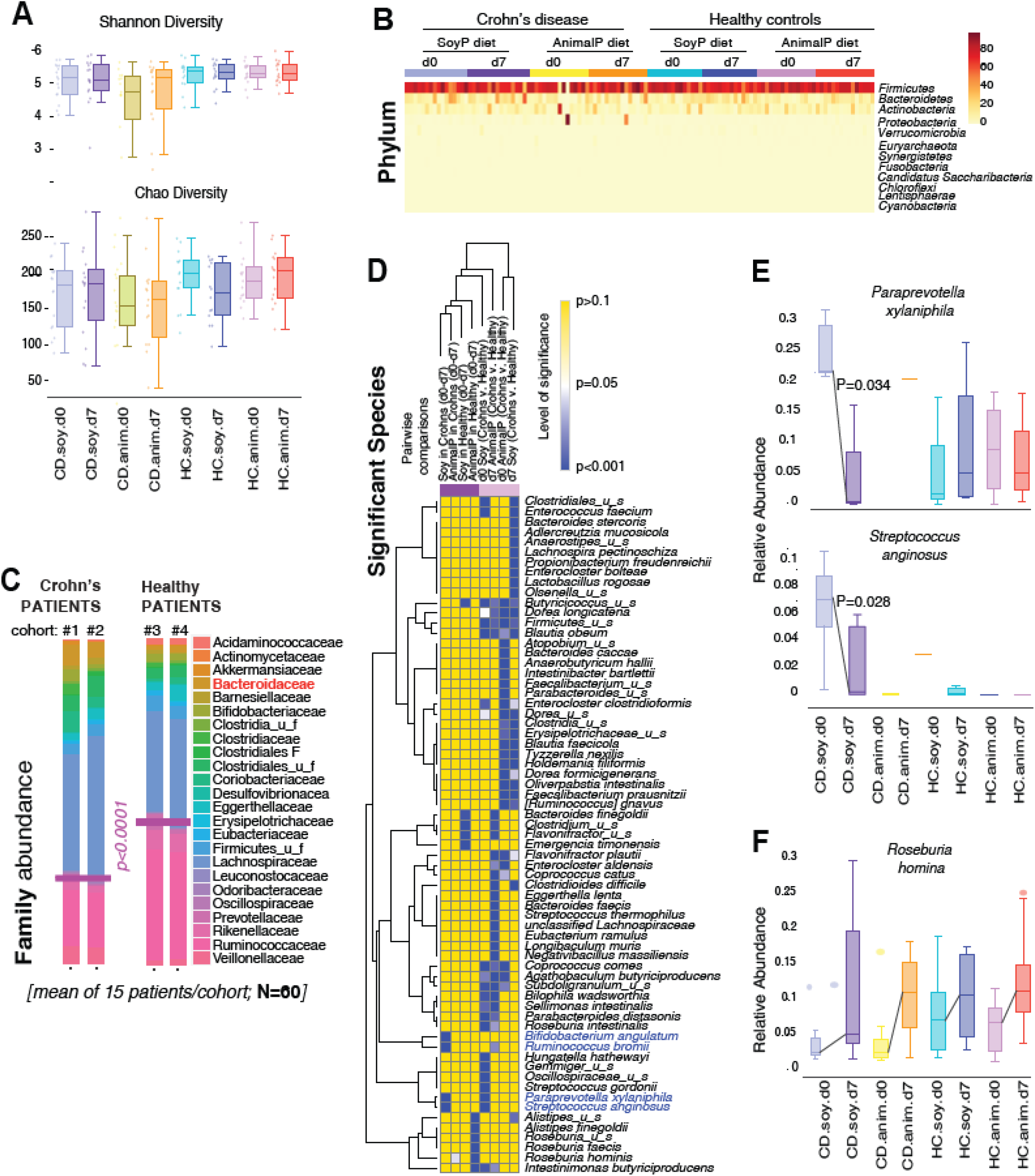
Microbiome composition and response to diet in Crohn’s disease and healthy individuals. **A)**. Alpha diversity indices, **B)**. Comparison of Phylum-level differences between CD and HC diet groups over time, **C)** Family-level comparison between CD and HC, **D)** Differential abundance analysis at the species level. **E-F)**. Comparison of select species between CD and HC diet groups.

Species-level analysis revealed 67 species that were significantly different across cohorts (representing approximately 67/806; 8.3% of the total number of species analyzed per group, **Figure 4D**). To control for inter-individual variability, we evaluated the relative change (d7-d0) in species abundance for each participant. Pairwise group analyses revealed that the CD SoyP-diet group had unique species shifts, including enrichment of *Roseburia hominis* and a reduction of *Paraprevotella xylaniphila* (*Prevotellaceae*) and *Streptococcus anginosus* (**Figure 4E-F**). Regarding *Streptococcus*, CD participants on the AnimalP-diet showed an increase in *Streptococcaceae* (P=0.045). While principal coordinate analysis (PCoA; Jaccard and Bray-Curtis dissimilarity) showed no substantial metagenomic segregation by diet type or disease status (**Figure 5A-B**) shifts in microbiome structure were observed at the individual-level (**Figure 5B**). Participant-level microbiome signature plots^25^, showed that the SoyP diet increased 23 putatively anti-inflammatory species while reducing 22 potentially pro-inflammatory species. (**Figure 5C**). This shift highlights the capacity of SoyP-diet to modulate species with potential anti-inflammatory effects in any particular individual, warranting long-term studies of diet-microbiome effects on CD immunoregulation.

**Figure 5.**
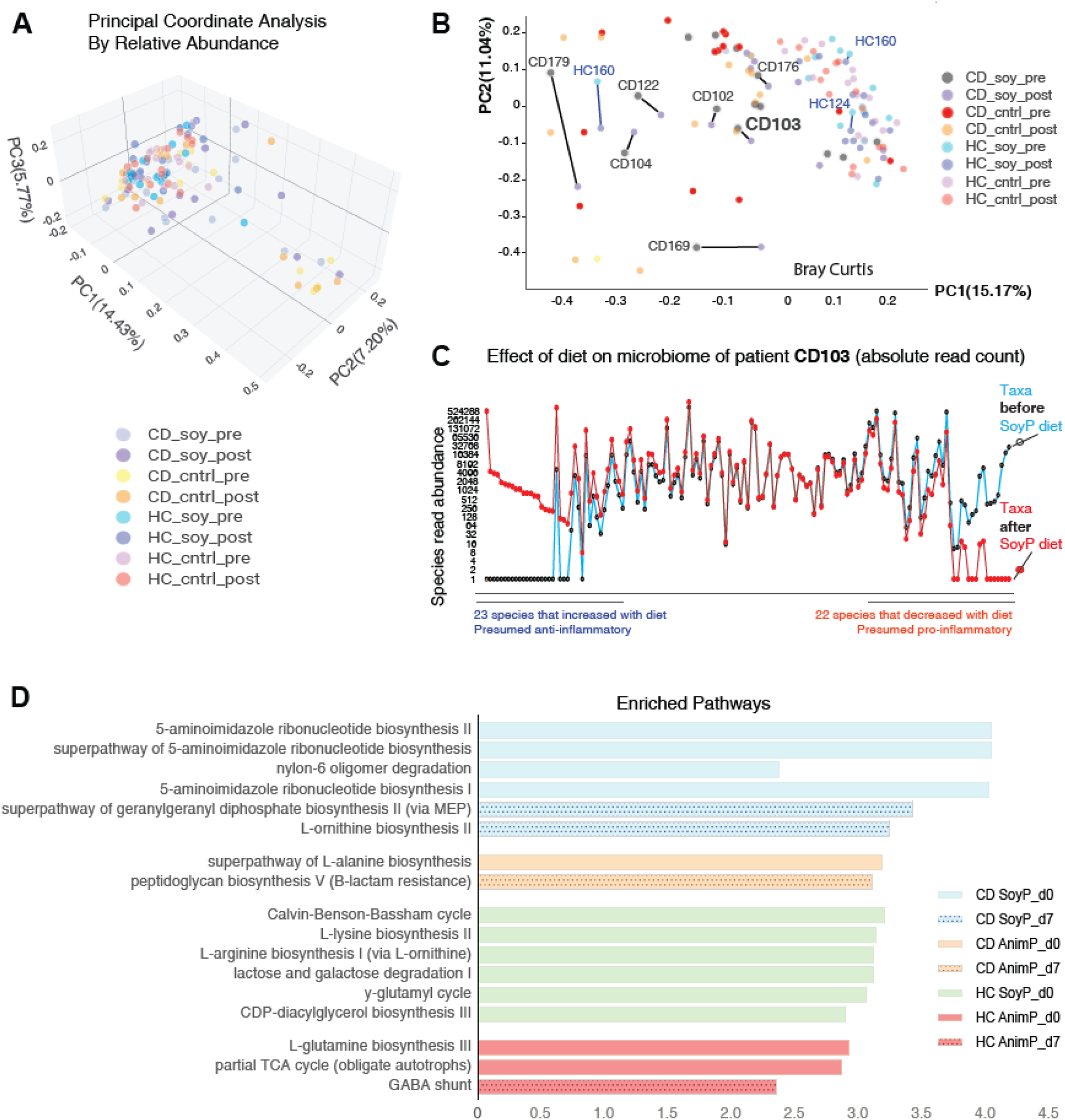
Microbiome and functional pathway shifts in response to diet. Principal coordinate analysis (PCoA) based on **A)** relative abundance data and **B)** Bray-Curtis dissimilarity, **C)** Individual-level paired signature line plot analysis of fecal microbiomes before and after dietary intervention, **D)** Microbial functional pathways (LeFse) that differed significantly between CD and HC diet groups, comparing day 0 to day 7

Linear discriminant analysis of effect size (LeFse) was used to identify functional pathways that differed significantly between CD and HC diet groups, comparing day 0 to day 7. Pathways differentially enriched between the CD and HC diet groups are shown in **Figure 5D**.

### Mice transplanted with CD-SoyP-diet human feces exhibit neutral or anti-inflammatory effects

We next examined whether the SoyP-diet-induced microbiota changes in CD participants could be transmissible to mice and lead to beneficial improvements using an acute DSS-colitis inflammation model. To do this, fecal microbiota transplantation (FMT) experiments were performed in germ-free SAMP mice using stool samples collected before and after the intervention (**Supplementary Figure 1A**).

The SoyP-diet exerted beneficial effects in all mice that received the CD SoyP-donor-transplanted feces (6/6 donors). Notably, improvements were observed in at least one of the parameters tested, although specific responses varied by donor. Although a slight increase in histology score was observed for one donor group, FMT transplants resulted primarily in anti-inflammatory effects across 28 mouse/parameter datasets (i.e., 6 donors and 4-5 variables assessed on mice/donor, pre-diet feces into 6 mice, vs post-diet feces into other 6 mice, see P-values in **Supplementary Figure 1B**). Collectively, mice transplanted with the SoyP-diet microbiota were more likely to exhibit a neutral effect (18/28) or a positive anti-inflammatory effect (9/28) due to the transplantation, rather than negative proinflammatory effects (1/28; P<0.00001; ordered pairwise Fisher’s exact comparisons P=0.0116 and P=0.0315). Since DSS was conducted after 3 weeks of transplantation, data indicate that the SoyP-diet may have transmissible lasting effects on the gut microbiota in humans that can be transferable to mice via FMT and exert consistently either neutral or positive anti-inflammatory effects in mice.

## DISCUSSION

This study is among the first RCTs to evaluate whether replacing animal protein with soy-pea protein in a controlled diet reduces the pro-inflammatory potential of the gut microbiota and intestinal inflammation in adults with clinically quiescent to moderate CD. The SoyP-diet was associated with significantly greater reductions in fecal MPO in 75% of CD participants, suggesting that short-term modification of dietary protein source, without restriction of other macronutrients, was associated with reductions in biochemical inflammation. These findings align with prospective cohort data showing that healthy plant-based dietary patterns are associated with a lower risk of IBD, including CD^12^. Although CDAI as a patient-reported outcome did not reach statistical significance, exploratory post-hoc composite analysis integrating changes in CDAI and fecal MPO showed greater improvement among participants assigned to the SoyP-diet compared with the AnimalP-diet. Consistent with this observation, other objective inflammatory biomarkers (FC, CRP) generally demonstrated directional changes favoring the SoyP-diet. Moreover, participants who completed both dietary arms had reproducible reductions in inflammatory markers during the SoyP-diet, supporting the observed anti-inflammatory effect. Overall, consumption of the SoyP-diet was safe, associated with few adverse events, and did not worsen disease activity.

This study included participants across a spectrum of disease activity. When stratified by baseline disease activity, the beneficial effects of the SoyP-diet were most pronounced among CD participants with CDAI <150, suggesting that patients who are clinically quiescent yet retain residual biochemical inflammation may derive the greatest benefit from SoyP dietary modification. Importantly, clinical remission in CD does not necessarily reflect absence of intestinal inflammation, and persistent subclinical inflammatory activity is associated with future relapse and disease progression^2, 3^. Therefore, even modest reductions in neutrophil-associated markers such as MPO may meaningfully influence disease trajectories. Accordingly, these findings should not be interpreted as evidence that the SoyP-diet induces remission in patients with highly active CD, but rather as support for a potential adjunctive role in reducing residual inflammatory burden during clinical remission. In contrast, participants with active disease (CDAI ≥150) demonstrated heterogeneous MPO responses, indicating that potential benefits in this subgroup may depend on additional, unmeasured modifiers.

Encouragingly, 66% of CD participants assigned to the SoyP-diet reported willingness to continue the intervention after food provision ended, indicating acceptability and potential for sustained use. Unlike restrictive approaches such as exclusive enteral nutrition, targeted modification of dietary protein may represent a more feasible strategy that integrates into existing eating patterns, consistent with evidence showing that palatability and sustainability are critical determinants of long-term adherence and clinical benefit.

While the effects of the SoyP-diet appeared largely independent of baseline microbiome composition at the group level, microbiome structure shifted within individuals following dietary intake, indicating that both SoyP- and AnimalP-diets elicit individualized changes in microbial community composition rather than uniform effects across participants (**Supplementary Discussion**). Notably, reductions in *Paraprevotella xylaniphila* (*Prevotellaceae* family), a species linked to CD through arabinoxylan and pectin metabolism and succinate production, were observed in the CD-SoyP group. Although succinate has been associated with microbiome dysbiosis and intestinal inflammation^26, 27^ its role in gut inflammatory processes remains incompletely defined. Functional analysis revealed CD-SoyP participants exhibited significant enrichment of L-ornithine biosynthesis and the superpathway of geranylgeranyl diphosphate biosynthesis II (via the MEP pathway), two pathways with established anti-inflammatory relevance to CD^28, 29^. Notably, microbiome-derived L-ornithine, has been associated with enhanced efficacy of biologic therapies such as ustekinumab by disrupting IL-23/STAT3 signaling in immune cells, boosting treatment efficacy^30^. In parallel, the SoyP-diet led to a significant decrease in 5-aminoimidazole ribonucleotide biosynthesis, a pathway which is part of the de novo purine synthesis pathway, shown to be enriched (upregulated) in CD inflammation^31-33^

Compositional differences inherent to plant-based protein sources may, in part, contribute to the observed microbiome and inflammatory shifts. In particular, soy-derived isoflavones were enriched in the SoyP diet and represent plausible bioactive mediators^34^. Collectively, these compounds have been shown to exert anti-inflammatory and antioxidant effects, improve intestinal barrier integrity, regulate key inflammatory pathways (e.g., NF-κB, p38), as well as beneficially modulate gut microbiota composition relevant to CD ^34-38^. Dietary iron intake was higher in the SoyP groups, although levels exceeded recommended daily allowances across all study groups. While higher luminal iron has been implicated in promoting intestinal inflammation and dysbiosis in CD^39^, the lower bioavailability of non-heme iron from plant sources suggests that the functional impact on intestinal inflammation may differ between diets ^40^. Although fiber and carbohydrate intake did not differ significantly between groups, similar directional trends for slightly higher intake in the SoyP groups were observed, suggesting that the intervention may reflect a broader shift in dietary composition rather than protein substitution alone.

This study has limitations, including a short 7-day intervention that precludes conclusions regarding long-term efficacy or disease trajectory effects. Accordingly, the findings should be interpreted as evidence of short-term biological responsiveness to dietary protein modification rather than definitive clinical efficacy or remission induction and require confirmation in longer-term studies. Second, providing all study foods enhanced internal validity but limited real-world generalizability. Exclusion of ultra-processed foods may have amplified SoyP effects, however, because all participants received comparable foods aside from protein type, this factor is unlikely to fully explain the observed differences in inflammatory markers. Third, the open-label design may have introduced expectation bias in patient-reported symptom outcomes (e.g., HBI, CDAI), and therefore symptom-based efficacy findings should be interpreted cautiously. However, inclusion of validated objective inflammatory biomarkers, including fecal MPO, FC, and hsCRP, as well as the subset of participants completing the study for a second time, helped mitigate reliance on symptom-based assessments alone. Moreover, concordant directional changes across multiple inflammatory markers, together with the differential response observed between CD and HC participants and supportive murine fecal microbiota transplantation findings, suggest that the observed effects are not solely attributable to day-to-day biological variability or placebo response. Fourth, objective evidence of active inflammation was not required for enrollment, and therefore some participants may have had functional gastrointestinal symptoms in the absence of active inflammatory disease. Nevertheless, many CD participants demonstrated biochemical evidence of ongoing inflammation at baseline, including elevated FC and MPO levels. Finally, despite randomization, baseline fecal MPO values were higher in the CD-SoyP group. Although adjusted analyses continued to support a significant diet-associated reduction in MPO it does not entirely eliminate the possibility of regression-to-the-mean effects. Future studies should incorporate objective inflammatory entry criteria and endoscopic endpoints to better define the subgroup most likely to benefit from dietary intervention.

In conclusion, this randomized controlled feeding study demonstrates that replacing animal protein with soy-pea protein in context of a controlled diet reduces biochemical inflammation in CD over a short period, with the greatest benefit observed in patients in clinical remission. These findings suggest that soy-pea may serve as a scalable, acceptable, and biologically meaningful adjunctive strategy for inflammation control and remission maintenance, and thus provide a strong rationale for longer-term RCTs evaluating whether sustained incorporation of soy-pea protein into habitual diets can modulate disease trajectory.

## Supporting information

CONSORT checklist

## Data Availability

Deidentified individual participant data are available indefinitely at figshare doi:10.6084/m9.figshare.30075568.

https://doi.org/10.6084/m9.figshare.30075568

## ABBREVIATIONS

AnimalP: animal protein
CD: Crohn’s disease
CDAI: Crohn’s Disease Activity Index
day 0: d0
day 7: d7
DHI: Digestive Health Institute
DSS: dextran sodium sulfate
EEN: exclusive enteral nutrition
GF: germ-free
HC: healthy control
HBI: Harvey Bradshaw Index
hsCRP: high-sensitivity C-reactive protein
IBD: inflammatory bowel disease
MPO: myeloperoxidase
SAMP: SAMP1/YitFC mouse
sCDAI: short Crohn’s disease activity index
SoyP: soy-pea protein
UC: ulcerative colitis
UHCMC: University Hospitals Cleveland Medical Center

## ACKNOWLEDGEMENTS

We gratefully acknowledge the participants who generously contributed their time and effort to this study. We also thank the members of the Data and Safety Monitoring Board for their oversight, guidance, and commitment to ensuring the integrity and safety of the trial.

## Notes

**FUNDING SUPPORT** Research reported in this publication was supported by an NIH/NIDDK F32DK117585 and K01DK127008, as well as by the Cleveland Digestive Diseases Research Core Center (DDRCC) Pilot Feasibility Award (NIH/NIDDK P30DK097948) (to ARB). Additional support of this work was provided by DK055812, DK091222, DK097948 and P01DK091222 Germ-free and Gut Microbiome Core (to FC), and R21DK118373 (to ARP). We want to acknowledge the support of the Mouse Models, the Histology Imaging, and Tissue Biorepository Cores of the NIH P30 Silvio O. Conte Cleveland Digestive Diseases Research Core Center.

**DISCLOSURES** Authors declare that there are no conflicts of interest or competing financial/non-financial interests to disclose. We confirm that this work is original and has not been published as peer-reviewed material elsewhere, nor is it currently under consideration for publication elsewhere. None of the authors or relatives have affiliations with the soy or pea or industry. Study was designed, analyzed, conducted and reported without external influence from any sector of the food industry.

### Competing Interest Statement

The authors have declared no competing interest.

### Clinical Trial

NCT04065048

### Author Declarations

The Institutional Review Board of University Hospitals Cleveland Medical Center gave ethical approval for this work (STUDY20190080).

### Summary of Updates

The revised manuscript includes additional analyses incorporating Bonferroni adjustment for multiple comparisons, as well as expanded discussion and clarification of study limitations.

