## Supplementary Materials for "A Randomized Controlled Trial Comparing Soy-Pea Protein to Animal Protein in Adults with Crohn’s Disease"

**Table of Contents**

**A. Supplementary Methods**

Assessment of Participants
Patient and Public Involvement
Adherence and Macronutrient Intake
Adverse Events
Protocol Deviations
Protocol Modifications
Monitoring and Data Quality
Laboratory Analysis
Fecal Microbiota Transplantation Experiments in Mice

**B. Supplementary Discussion**

**C. Supplementary Tables**

**Supplementary Table 1.** Self-reported Foods Consumed by Participants on the Soy-pea Protein Diet that were Not Provided by the Study
**Supplementary Table 2.** Self-reported Foods Consumed by Participants on the Animal Protein Diet that were Not Provided by the Study
**Supplementary Table 3.** Within-group Total Daily Macronutrient and Fiber Intake Among Males
**Supplementary Table 4.** Within-group Total Daily Macronutrient and Fiber Intake Among Females
**Supplementary Table 5.** Between-group Total Daily Macronutrient and Fiber Intake Among Males
**Supplementary Table 6.** Between-group Total Daily Macronutrient and Fiber Intake Among Females
**Supplementary Table 7.** Within-group Total Micronutrient Intake Over 7 Days Among Male Crohn's Disease Participants by Diet (SoyP vs AnimalP)
**Supplementary Table 8.** Within-group Total Micronutrient Intake Over 7 Days Among Male Healthy Control Participants by Diet (SoyP vs AnimalP)
**Supplementary Table 9.** Within-group Total Micronutrient Intake Over 7 Days Among Female Crohn's Disease Participants by Diet (SoyP vs AnimalP)
**Supplementary Table 10.** Within-group Total Micronutrient Intake Over 7 Days Among Female Healthy Control Participants by Diet (SoyP vs AnimalP)
**Supplementary Table 11.** Between-group Total Micronutrient Intake Over 7 Days Among Male Crohn's Disease and Healthy Control Participants Assigned the SoyP diet
**Supplementary Table 12**. Between-group Total Micronutrient Intake Over 7 Days Among Male Crohn's Disease and Healthy Control Participants Assigned the AnimalP diet
**Supplementary Table 13.** Between-group Total Micronutrient Intake Over 7 Days Among Female Crohn's Disease and Healthy Control Participants Assigned the SoyP diet
**Supplementary Table 14.** Between-group Total Micronutrient Intake Over 7 Days Among Female Crohn's Disease and Healthy Control Participants Assigned the AnimalP diet

**D. Supplementary Figure**

**Supplementary Figure 1.** Fecal microbiota transplantation experiments using baseline ("pre") and diet-altered ("post") stool collected from Crohn's disease participants assigned to the SoyP-diet intervention.

**E. Supplementary References**

### A. Supplementary Methods

#### *Assessment of Participants*

We used a combination of participant self-reported outcomes and medical records for demographic information, CD history, medical and medication history, dietary recalls, and disease symptoms. Disease activity was measured using the Crohn's Disease Activity Index (CDAI, primary outcome) and HBI. Fecal MPO, fecal calprotectin (FC), and high-sensitivity C-reactive protein (hsCRP) were measured to assess the inflammatory response to the diet. Fecal calprotectin (FC) analysis was conducted in a single batch at the conclusion of the study. Final analyses of FC data excluded one CD AnimalP-diet participant due to an erroneous value quantified by the kit.

Our group has demonstrated in animal models and humans that fecal MPO changes in response to diet[1], and represents an optimal marker to assess the inflammatory response to diet. In the present study, we used fecal MPO as a surrogate marker to evaluate changes in intestinal inflammation in CD patients following the dietary intervention. We and others have shown that the presence of neutrophils in feces, quantified by MPO activity (a primary constituent of neutrophils) and by fecal calprotectin (FC), serve as a reliable indicator for monitoring intestinal inflammation in IBD.

#### *Patient and Public Involvement*

Patients and the public were not involved in the development of the research question, outcome measures, or study design. The research questions and endpoints were informed by prior preclinical and translational work. Participants were first involved at the point of recruitment and participation in the study. Although patients were not involved in the design or conduct of the study, efforts were made to minimize participant burden. At the conclusion of the study, participants completed a diet satisfaction survey to assess acceptability and perceived burden of the intervention. Study findings will be disseminated to participants through a lay-language summary of the results, and efforts will be made to share findings with relevant patient communities in accessible formats. Future studies will incorporate patient and public involvement in the design and refinement of research questions, outcome measures, and dissemination strategies to better align with patient priorities and preferences.

#### *Adherence and Macronutrient Intake*

Foods that were consumed but which were not provided by the study diet were determined as either ‘compliant’ or ‘non-compliant’ based on the food type. For example, the consumption of cow’s milk (if not provided by the study) by a participant on the AnimalP-diet would be deemed ‘compliant’, but deemed as ‘non-compliant’ for a participant on the SoyP-diet. Overall, only one participant on the SoyP-diet reported a single occurrence of consuming a non-compliant food item (skim milk). Foods consumed that were not provided by the study are shown in **Supplementary Tables 1-2.**

#### *Adverse Events*

Adverse events were assessed at end of study visit (day 7 of the study diet). Serious adverse events included any adverse event that was fatal, life-threatening, requiring prolonged hospital stay, congenital anomaly or birth defect or other medically significant event as deemed such by the investigator.

Overall, most of the adverse events reported by participants at the end of study visit were gastrointestinal related, and resolved within the first 1-2 days of the study diet. Bloating/flatulence was the most common complaint reported (SoyP diet (CD; 12.5%, 2/16, HC; 28.5%, 4/14) vs AnimalP (CD; 20%, 3/15], followed by constipation (SoyP; CD 6.25%, 1/16) and increased stool (SoyP; CD 6.25%, 1/16). Headache was reported by one CD SoyP and one CD AnimalP participant.

A total of two adverse events that were possibly related to the study diet were reported (attrition rate of 6.6%, n=2/30 participants in the SoyP-diet groups; 1 CD and 1 healthy control participant). In brief, one female Crohn’s disease (CD) participant assigned to the soy-based diet experienced abdominal discomfort and nausea in the evening of day 1 after starting the soy-based diet. The participant was instructed to immediately stop the study diet and symptoms resolved within 48 hours. The second additional adverse event was reported in one female HC participant assigned to the soy-based diet who reported experiencing a headache and nausea after starting the study diet. However, the participant reported “I believe I experienced a headache, low levels of energy and nausea as a result of not consuming enough calories during the day and not receiving my normal protein intake”. No other adverse events were reported. No Deaths or other Unanticipated problems were reported.

***Protocol Deviations***

Protocol deviations that occurred early in the study included: (1) Improper completion of the nurse flow sheet during visit 1 for one participant, (2) use of an outdated version of the consent form during the consenting process for six participants, and (3) initiation of the study diet more than 14 days after stool sample 1 was collected for one participant. In addition, although the protocol specified an HBI score <4 or >8, CD participants with scores between 5-7 were included. None of these deviations impacted the completeness, accuracy, or reliability of the study data.

***Protocol Modifications***

This study began enrolling in September 2019, just before the onset of the COVID-19 pandemic. Due to health mandates implemented during the pandemic, study activities were temporarily halted. To ensure the continuation of the study while minimizing the burden of in-person hospital visits, the following amendments were made to the study protocol: (1) Consent and study procedures, initially planned as in-person, were conducted remotely via Doxy.me, following the "Guidelines for Remote Electronic Consent" provided by University Hospitals Cleveland Medical Center, and (2) Blood draws were arranged to take place at participants' homes through the local home health care company BrightStar Care.

The original study was developed based on a 2019 Cleveland Digestive Diseases Research Core Center (DDRCC) pilot feasibility award (P30DK097948) and initially focused exclusively on individuals with Crohn's disease (CD). However, in 2020, the principal investigator (ARB) secured additional funding (K01DK127008-01), which enabled the expansion of the study to include healthy controls (HC) and an increase in the proposed sample size from 12 CD participants to 60 subjects (30 CD and 30 HC). Additionally, fecal myeloperoxidase (MPO) activity was incorporated as a primary outcome to allow comparisons between Crohn's patients and controls. The quantification of fecal MPO had already been approved in the original protocol. Initially, the study's inclusion criteria required participants to be between 18 and 55 years old. After five months of recruitment, the study steering committee expanded the age range to 18-65 years to broaden eligibility. Furthermore, an additional gastroenterologist was added to the study as an investigator to support expanded patient recruitment efforts.

***Monitoring and Data Quality***

This study was overseen by a Data Safety Monitoring Board (DSMB), which convened at three key points: prior to the study's initiation, after half of the participants were enrolled, and at the conclusion of the trial. The monitoring focused on regulatory documentation, the consent process, participant eligibility and interim analyses of data. DSMB meetings followed a standard format, starting with an open session involving both DSMB members and study investigators. This was followed by a closed session where unblinded data could be reviewed, and, if necessary, an additional open session was held. In December 2022, the study underwent a comprehensive research compliance audit conducted by University Hospitals. The audit found the study to be in good standing, with only minor compliance issues noted. All required actions from the audit's final report were addressed, resolved, and documented in the study's regulatory binder.

***Laboratory Analysis***

Blood serum was collected and analyzed for hematocrit and high-sensitivity C-reactive protein (hsCRP) by University Hospitals Laboratory Services. Freshly voided stool samples were stored at -80°C within 2 hours of collection. Fecal MPO activity was assessed (measured in triplicate) within 5 days of collection, as previously described[2, 3].

***Fecal Microbiota Transplantation Experiments in Mice***

To understand the role of microbiome changes on inflammation, we conducted a series of fecal microbiota experiments. Groups of six 14-week-old sex-matched (littermate controls) inbred germ-free (GF) SAMP1/YitFC (SAMP) mice (Cleveland Digestive Diseases Research Center, Mouse Models Core) transplanted with either 'pre' (d0) or 'post' (d7) SoyP or AnimalP diet-altered human feces were used to quantify the inflammatory (functional) potential diet-altered human gut microbiota. Details on the human gut microbiota transplanted GF SAMP (hGM-SAMP) model is previously described[4]. Two weeks post colonization, colitis was induced with 3% DSS (TdB Consultancy AB) offered ad libitum for 7 days and mice resumed with water for 2 days, then were killed. The impact of FMT was assessed using five parameters associated with colonic inflammation (MPO, FITC, colon length, and histology).

We have demonstrated that germ-free (GF) SAMP mice transplanted with human gut microbiota (hGM-SAMP) and treated with dextran sodium sulfate; DSS (3% for 7days) display increased susceptibility to DSS-associated morbidity depending on the 'pro-inflammatory' potential of human donor feces[4, 5]. Concurrent reports support temporal differences in the IBD microbiome[6]. We used our validated hGM-SAMP DSS model as a model of gut barrier dysfunction (described in detail[4, 5]) to quantify the inflammatory (functional) potential of SoyP and AnimalP diet-altered human gut microbiota. Fecal myeloperoxidase (MPO) activity was assessed in mouse feces (measured in triplicate) as described[3, 7]. Intestinal permeability was measured using a fluorescently labeled small molecule [fluorescein isothiocyanate (FITC)-dextran], as described[8]. Colonic tissue samples were submitted for blinded histological assessment of formalin-fixed tissues using validated methodology[2, 9]. Murine colonoscopy was performed on the day of sacrifice using a flexible digital ureteroscope (URF-V; Olympus America) and inflammation quantified using a validated endoscopic scoring system[10].

Animals were maintained on a standard rodent chow following weaning. Importantly, animals were not fed soy- or animal-protein diets resembling the human interventions, in order to determine whether the hypothesized microbiome-mediated anti-inflammatory effects observed in humans could also be detected in mice when maintained on a different diet. Thus, throughout the experiment, all mice were fed laboratory rodent pellets, standard to our facility (Labdiet® Rat/Mouse 18%VacPac-5LQ6, Charles River), and which have been used in our facility to characterize the SAMP mouse CD-like ileitis phenotype in various studies[2, 7, 11]. All diets were vacuum packed and double irradiated to reach GF standards. All mice were individually housed on nonedible Aspen bedding in our GF-grade NesTiso caging system[12] and maintained on a 12-h:12-h light:dark cycle in a species-appropriate, temperature- and humidity-controlled room within AAALAC-accredited Animal Research Center facilities at Case Western Reserve University. All procedures were approved by the Institutional Animal Care and Use Committee and the Institutional Review Board at CWRU, in accordance with the Guide for Care and Use of Laboratory Animals.

### B. Supplementary Discussion

*Effect of the SoyP diet on fecal microbiome*

While the effects of the SoyP diet appeared largely independent of baseline microbiome composition at the group level, microbiome structure shifted within individuals following dietary intake, indicating that both SoyP and AnimalP diets elicit modest individualized, rather than uniform, changes in microbial community composition. Notably, reductions in *Paraprevotella xylaniphila* (*Prevotellaceae* family), a species linked to CD through arabinoxylan and pectin metabolism and succinate production, were observed in the CD SoyP diet group. Although succinate has been associated with microbiome dysbiosis and intestinal inflammation[13, 14] its role in gut inflammatory processes remains incompletely defined.

Although *Enterobacteriaceae* were detected, they were not prominent in this dataset and were not among the taxa most influenced by diet. The increased abundance of *Streptococcaceae* observed among the CD AnimalP-diet group is of particular interest however, as the role of *Streptococcaceae* in CD remains poorly understood. The genus *Streptococcus* includes multiple species that differ in their pathogenic potential due to the presence of strain-specific toxins and virulence factors. Notably, alterations in *Streptococcus* abundance (*S. anginosus*) were also observed, albeit in the opposite direction in CD individuals consuming the SoyP-diet. This contrasting pattern suggests a diet-dependent modulation of *Streptococcus* populations, which may warrant further investigation in future studies.

Functional analysis (Lefse) revealed that, among CD participants, 7 days of the SoyP diet resulted in significant enrichment of two pathways with established anti-inflammatory relevance to CD, namely L-ornithine biosynthesis and the superpathway of geranylgeranyl diphosphate biosynthesis II (via the MEP pathway)[15, 16]. Microbiome-derived L-ornithine, produced by beneficial taxa such as *Faecalibacterium prausnitzii*, has been associated with enhanced efficacy of biologic therapies such as ustekinumab by disrupting IL-23/STAT3 signaling in immune cells, boosting treatment efficacy[17]. In this context, *F. prausnitzii* abundance increased in 37.5% (6/16) of CD participants consuming the SoyP-diet (paired t-test, P = 0.33). Enrichment of the geranylgeranyl diphosphate biosynthesis pathway has been associated with anti-inflammatory effects in experimental models of colitis, in part through its role in the maintenance and differentiation of regulatory T cells (Tregs)[16]. In parallel, the SoyP diet led to a significant decrease in 5-aminoimidazole ribonucleotide biosynthesis, a pathway which is part of the de novo purine synthesis pathway, shown to be enriched (upregulated) in CD inflammation[18-20]. Specifically, purine metabolism has been linked to intestinal permeability and regulating immune response in inflamed intestinal tissue (e.g., LC3/T helper type 17-interleukin 22 pathway)[21]. Collectively, these findings support the concept that a SoyP diet may serve to enhance or prolong remission response to traditional IBD therapies. By comparison, AnimalP intake led to an enrichment of peptidoglycan biosynthesis V (β-lactam resistance) with reductions in L-alanine biosynthesis, consistent with prior reports linking bacterial cell wall metabolism, NOD2 signaling, and immune activation to CD pathogenesis[22, 23]. Among HC participants, AnimalP intake was associated with enrichment of the GABA shunt and reductions in pathways related to L-glutamine biosynthesis. Although pathways such as the Calvin-Benson-

Bassham cycle and partial TCA variants do not directly correspond to human metabolism, they have been reported as microbial functional markers distinguishing health, remission, and therapeutic response states in IBD[24].

### C. Supplementary Tables

**Supplementary Table 1.** Self-reported Foods Consumed by Participants on the Soy-pea Protein Diet that were Not Provided by the Study

| Compliance N=16* | Foods |
|---|---|
| Compliant foods consumed that were not provided by the study, n=15 (94%) | fruit cup, mango juice, bell pepper, lemon, lime, Brussel sprouts, coconut, Brazil nuts, soy milk, romaine lettuce, mushrooms, pickles, hot sauce, apple, banana, celery, lime juice, rice vinegar, cupcake, granola, cookie, 5 cinnamon mints |
| Non-compliant foods consumed that were not provided by the study, n=1 (6%) | glass of skim milk |

*A total of 16 of the 30 participants on the soy diet reported consuming foods that were not provided by the study, of whom 15 (50%) had foods that were still considered 'compliant' to the diet, and one participant reporting a single occurrence of consuming a non-compliant food.

**Supplementary Table 2.** Self-reported Foods Consumed by Participants on the Animal Protein Diet that were Not Provided by the Study

| Compliance N=17* | Foods |
| --- | --- |
| Compliant foods consumed that were not provided by the study, n=17 (100%) | cheese, lemon, kiwi, apple, fruit cup, cucumber, avocado, broccoli sprouts, steak, chicken, broccoli, hamburger, lettuce, pickles, steak, soy free sauce, turkey bacon, butter, pepperoni, balsamic vinegar, stir fry veg, onions, celery, frozen veg, crackers, doughnut, breaded chicken wing, hard candy |
| Non-compliant foods consumed that were not provided by the study | n/a |

*A total of 17 of the 30 participants on the animal protein diet reported consuming foods that were not provided by the study, of which all participants (100%) had foods that were still considered ‘compliant’ to the diet.

**Supplementary Table 3.** Within-group Total Daily Macronutrient and Fiber Intake Among Males

| | Crohn's Disease | Crohn's Disease | |
|---|---|---|---|
| | SoyP (n=8) | AnimalP (n=5) | P-value* |
| Kcals | 1894.8 (308.4) | 1918.5 (443.8) | 0.911 |
| Fat (g) | 90.7 (24.2) | 108.6 (28.2) | 0.248 |
| Carb (g) | 203 (48.3) | 150.6 (48.8) | 0.085 |
| Pro (g) | 102.8 (18.2) | 105.3 (16.4) | 0.808 |
| Total Fiber | 34.6 (8.1) | 22.2 (6.7) | 0.017 |
| | Healthy Control | Healthy Control | |
| | SoyP (n=3) | AnimalP (n=3) | P-value* |
| Kcals | 2233.5 (828.5) | 1745.5 (212.6) | 0.183 |
| Fat (g) | 105.6 (57.6) | 89.4 (14.9) | 0.661 |
| Carb (g) | 276.5 (108) | 171.2 (31.7) | 0.181 |
| Pro (g) | 128.36 (40.88) | 86.9 (13.1) | 0.170 |
| Total Fiber | 50.3 (24.2) | 19.6 (4.8) | 0.098 |

AnimalP; animal protein diet, SoyP; soy-pea protein diet.

*Unpaired t-test p. Calculations include self-reported foods not provided by the study. Dietary intake data collected and analyzed using Nutrition Data System for Research software version 2019, developed by the Nutrition Coordinating Center (NCC), University of Minnesota, Minneapolis, MN. Results are mean, standard deviation.

**Supplementary Table 4.** Within-group Total Daily Macronutrient and Fiber Intake Among Females

| | Crohn's Disease | Crohn's Disease | |
|---|---|---|---|
| | SoyP (n=8) | AnimalP (n=10) | P-value* |
| Kcals | 1826.5 (246.8) | 1761.7 (394.6) | 0.691 |
| Fat (g) | 95.4 (24.4) | 92.7 (26.2) | 0.826 |
| Carb (g) | 199.4 (43.6) | 167.2 (39.6) | 0.121 |
| Pro (g) | 78.0 (8.9) | 87.9 (19.9) | 0.212 |
| Total Fiber | 31.1 (5.8) | 26.9 (12.80) | 0.415 |
| | Healthy Control | Healthy Control | |
| | SoyP (n=11) | AnimalP (n=12) | P-value* |
| Kcals | 1935.6 (691.0) | 2050.8 (413.6) | 0.629 |
| Fat (g) | 103.2 (43.6) | 119.6 (29.1) | 0.297 |
| Carb (g) | 205.1 (68.0) | 171.2 (40.5) | 0.157 |
| Pro (g) | 83.0 (30.1) | 97.9 (23.1) | 0.195 |
| Total Fiber | 35.1 (16.17) | 26.9 (4.82) | 0.109 |

AnimalP; animal protein diet, SoyP; soy-pea protein diet.

*Unpaired t-test p. Calculations include self-reported foods not provided by the study. Dietary intake data collected and analyzed using Nutrition Data System for Research software version 2019, developed by the Nutrition Coordinating Center (NCC), University of Minnesota, Minneapolis, MN. Results are mean, standard deviation.

**Supplementary Table 5.** Between-group Total Daily Macronutrient and Fiber Intake Among Males

| | Healthy Control | Crohn's Disease | |
|---|---|---|---|
| | SoyP (n=3) | SoyP (n=8) | P-value* |
| Kcals | 2233.5 (828.5) | 1894.8 (308.4) | 0.320 |
| Fat (g) | 105.6 (57.6) | 90.7 (24.2) | 0.661 |
| Carb (g) | 276.5 (108) | 203 (48.3) | 0.136 |
| Pro (g) | 128.36 (40.88) | 102.8 (18.2) | 0.166 |
| Total Fiber | 50.3 (24.2) | 34.6 (8.1) | 0.101 |
| | AnimalP (n=3) | AnimalP (n=5) | P-value* |
| Kcals | 1745.5 (212.6) | 1918.5 (443.8) | 0.558 |
| Fat (g) | 89.4 (14.9) | 108.6 (28.2) | 0.325 |
| Carb (g) | 171.2 (31.7) | 150.6 (48.8) | 0.543 |
| Pro (g) | 86.9 (13.1) | 105.3 (16.4) | 0.152 |
| Total Fiber | 19.6 (4.82) | 22.2 (6.7) | 0.601 |

AnimalP; animal protein diet, SoyP; soy-pea protein diet.

*Unpaired t-test p. Calculations include self-reported foods not provided by the study. Dietary intake data collected and analyzed using Nutrition Data System for Research software version 2019, developed by the Nutrition Coordinating Center (NCC), University of Minnesota, Minneapolis, MN. Results are mean, standard deviation.

**Supplementary Table 6.** Between-group Total Daily Macronutrient and Fiber Intake Among Females

| | Healthy Control | Crohn's Disease | |
|---|---|---|---|
| | SoyP (n=11) | SoyP (n=8) | P-value* |
| Kcals | 1935.6 (691.0) | 1826.5 (246.8) | 0.676 |
| Fat (g) | 103.2 (43.6) | 95.4 (24.4) | 0.655 |
| Carb (g) | 205.1 (68.0) | 199.4 (43.6) | 0.838 |
| Pro (g) | 83.0 (30.1) | 78.0 (8.9) | 0.656 |
| Total Fiber | 35.1 (16.1) | 31.1 (5.8) | 0.510 |
| | AnimalP (n=12) | AnimalP (n=10) | P-value* |
| Kcals | 2050.8 (413.6) | 1761.7 (394.6) | 0.111 |
| Fat (g) | 119.6 (29.1) | 92.7 (26.2) | 0.035 |
| Carb (g) | 171.2 (40.5) | 167.2 (39.6) | 0.818 |
| Pro (g) | 97.9 (23.1) | 87.9 (19.9) | 0.295 |
| Total Fiber | 26.9 (4.8) | 26.9 (12.8) | 0.939 |

AnimalP; animal protein diet, SoyP; soy-pea protein diet.

*Unpaired t-test p. Calculations include self-reported foods not provided by the study. Dietary intake data collected and analyzed using Nutrition Data System for Research software version 2019, developed by the Nutrition Coordinating Center (NCC), University of Minnesota, Minneapolis, MN. Results are mean, standard deviation.

**Supplementary Table 7.** Within-group Total Micronutrient Intake Over 7 Days Among Male Crohn's Disease Participants by Diet (SoyP vs AnimalP)

| | SoyP (n=8) | AnimalP (n=5) | P-value* |
|---|---|---|---|
| Daidzein (mg) | 332.95 (101.10) | 0.69 (0.94) | 0.00002 |
| Genistein (mg) | 428.83 (137.15) | 1.29 (1.59) | 0.00003 |
| Glycitein (mg) | 79.88 (26.43) | 0.13 (0.26) | 0.00004 |
| Coumestrol (mg) | 0.37 (0.38) | 0.19 (0.23) | 0.379 |
| Biochanin A (mg) | 0.74 (2.22) | 2.44 (4.79) | 0.399 |
| Formononetin (mg) | 0.07 (0.03) | 0.06 (0.03) | 0.648 |
| Tryptophan (g) | 7.73 (1.50) | 7.70 (1.43) | 0.973 |
| Threonine (g) | 22.65 (4.00) | 26.51 (4.20) | 0.124 |
| Isoleucine (g) | 26.38 (5.38) | 29.46 (4.64) | 0.313 |
| Leucine (g) | 44.74 (8.99) | 49.33 (8.81) | 0.387 |
| Lysine (g) | 33.97 (7.02) | 43.69 (7.60) | 0.038 |
| Methionine (g) | 8.18 (1.54) | 14.66 (2.31) | 0.0001 |
| Cystine (g) | 7.98 (1.41) | 8.28 (2.04) | 0.764 |
| Phenylalanine (g) | 29.91 (6.00) | 28.26 (5.29) | 0.624 |
| Tyrosine (g) | 20.69 (4.10) | 22.94 (3.31) | 0.327 |
| Valine (g) | 28.67 (5.28) | 33.32 (6.39) | 0.180 |
| Arginine (g) | 49.87 (9.52) | 40.99 (12.03) | 0.166 |
| Histidine (g) | 15.58 (3.13) | 17.42 (3.15) | 0.325 |
| Alanine (g) | 26.27 (4.79) | 30.60 (5.86) | 0.172 |
| Aspartic Acid (g) | 70.80 (14.09) | 60.57 (11.28) | 0.199 |
| Glutamic Acid (g) | 121.34 (21.95) | 113.88 (23.57) | 0.573 |
| Glycine (g) | 26.99 (4.84) | 27.80 (6.90) | 0.806 |
| Proline (g) | 31.79 (6.65) | 35.75 (6.17) | 0.306 |
| Serine (g) | 31.95 (6.58) | 29.53 (6.05) | 0.521 |
| Vit A (IU) | 64184.54 (31563.76) | 101527.99 (25105.71) | 0.048 |
| Vit D (mcg) | 29.55 (8.54) | 24.86 (9.41) | 0.374 |
| Vit E (mg) | 68.64 (22.18) | 78.11 (29.66) | 0.523 |
| Vit K (mcg) | 1680.65 (622.16) | 1581.29 (299.90) | 0.748 |
| Vit C (mg) | 1215.37 (646.61) | 929.65 (310.65) | 0.381 |
| Thiamin (mg) | 8.92 (2.43) | 5.45 (2.40) | 0.029 |
| Riboflavin (mg) | 11.12 (2.09) | 11.57 (1.65) | 0.691 |
| Niacin (mg) | 94.01 (35.69) | 152.32 (42.68) | 0.022 |
| Pantothenic Acid (mg) | 28.95 (10.22) | 40.14 (9.37) | 0.073 |
| Vit B-6 (mg) | 12.37 (3.21) | 14.34 (3.86) | 0.338 |
| Folate (mcg) | 3710.79 (83.53) | 2078.02 (355.06) | <0.000001 |
| B- 12 (mcg) | 3710.79 (8.38) | 29.71 (6.10) | <0.000001 |
| Calcium (mg) | 6351.26 (1053.21) | 6709.49 (1248.65) | 0.589 |
| Phosphorous (mg) | 9198.60 (1622.75) | 9564.23 (2398.35) | 0.747 |
| Magnesium (mg) | 3430.84 (861.19) | 2494.85 (912.66) | 0.089 |
| Iron (mg) | 135.19 (24.01) | 77.29 (28.74) | 0.002 |
| Zinc (mg) | 65.87 (15.39) | 83.77 (28.15) | 0.162 |
| Copper (mg) | 23.14 (22.74) | 11.98 (6.74) | 0.315 |
| Selenium (mcg) | 244.48 (71.65) | 615.81 (156.47) | 0.0001 |
| Sodium (mg) | 16647.12 (9780.58) | 17421.38 (5389.14) | 0.875 |
| Potassium (mg) | 28267.74 (5368.04) | 21801.23 (5726.76) | 0.064 |

AnimalP; animal protein diet, SoyP; soy-pea protein diet.

*Unpaired t-test p. Calculations include self-reported foods not provided by the study. Dietary intake data collected and analyzed using Nutrition Data System for Research software version 2019, developed by the Nutrition Coordinating Center (NCC), University of Minnesota, Minneapolis, MN. Results are mean, standard deviation.

**Supplementary Table 8.** Within-group Total Micronutrient Intake Over 7 Days Among Male Healthy Control Participants by Diet (SoyP vs AnimalP)

| | SoyP (n=3) | AnimalP (n=3) | P-value* |
|---|---|---|---|
| Daidzein (mg) | 331.28 (87.95) | 0.12 (0.07) | 0.003 |
| Genistein (mg) | 434.28 (115.04) | 0.72 (0.68) | 0.003 |
| Glycitein (mg) | 66.63 (17.11) | 0.00 (0.00) | 0.003 |
| Coumestrol (mg) | 0.26 (0.23) | 0.04 (0.07) | 0.178 |
| Biochanin A (mg) | 0.05 (0.01) | 0.02 (0.02) | 0.081 |
| Formononetin (mg) | 0.14 (0.02) | 0.04 (0.02) | 0.002 |
| Tryptophan (g) | 9.64 (2.86) | 6.39 (1.25) | 0.146 |
| Threonine (g) | 27.87 (8.54) | 21.05 (4.04) | 0.280 |
| Isoleucine (g) | 33.08 (10.12) | 23.83 (4.67) | 0.224 |
| Leucine (g) | 55.87 (17.30) | 40.58 (7.25) | 0.231 |
| Lysine (g) | 41.10 (12.59) | 35.96 (7.00) | 0.569 |
| Methionine (g) | 10.56 (3.10) | 11.63 (2.39) | 0.659 |
| Cystine (g) | 10.53 (3.09) | 6.71 (0.90) | 0.109 |
| Phenylalanine (g) | 38.12 (12.23) | 23.28 (4.56) | 0.120 |
| Tyrosine (g) | 25.11 (7.87) | 18.10 (4.27) | 0.247 |
| Valine (g) | 36.56 (10.99) | 27.03 (5.28) | 0.248 |
| Arginine (g) | 64.47 (20.33) | 36.42 (4.42) | 0.080 |
| Histidine (g) | 19.66 (6.32) | 14.52 (2.36) | 0.257 |
| Alanine (g) | 33.12 (10.61) | 25.24 (4.15) | 0.297 |
| Aspartic Acid (g) | 88.42 (29.18) | 52.08 (10.84) | 0.113 |
| Glutamic Acid (g) | 156.08 (49.12) | 97.25 (18.35) | 0.124 |
| Glycine (g) | 35.79 (11.99) | 22.99 (2.68) | 0.145 |
| Proline (g) | 38.69 (12.41) | 27.49 (7.16) | 0.247 |
| Serine (g) | 39.52 (12.28) | 24.26 (4.65) | 0.114 |
| Vit A (IU) | 46437.68 (17985.98) | 25606.07 (20788.15) | 0.260 |
| Vit D (mcg) | 32.09 (7.29) | 39.33 (51.87) | 0.823 |
| Vit E (mg) | 175.24 (116.50) | 73.31 (18.16) | 0.209 |
| Vit K (mcg) | 2078.73 (1405.88) | 1147.01 (1114.83) | 0.419 |
| Vit C (mg) | 1520.68 (786.49) | 873.48 (873.09) | 0.394 |
| Thiamin (mg) | 9.85 (3.01) | 4.61 (0.77) | 0.043 |
| Riboflavin (mg) | 13.23 (5.75) | 9.65 (3.67) | 0.416 |
| Niacin (mg) | 124.37 (50.67) | 127.40 (37.03) | 0.937 |
| Pantothenic Acid (mg) | 48.12 (20.12) | 33.16 (12.99) | 0.340 |
| Vit B-6 (mg) | 18.04 (6.39) | 12.27 (3.86) | 0.251 |
| Folate (mcg) | 4160.44 (1649.38) | 1520.37 (1003.23) | 0.077 |
| B- 12 (mcg) | 24.93 (5.44) | 20.10 (7.79) | 0.428 |
| Calcium (mg) | 7095.85 (3053.84) | 4054.93 (1727.81) | 0.208 |
| Phosphorous (mg) | 13327.84 (3774.09) | 8144.98 (1688.50) | 0.096 |
| Magnesium (mg) | 4741.40 (1567.74) | 2496.95 (237.06) | 0.070 |
| Iron (mg) | 167.75 (54.40) | 63.10 (5.24) | 0.029 |
| Zinc (mg) | 94.80 (27.10) | 65.57 (11.15) | 0.159 |
| Copper (mg) | 32.94 (9.12) | 11.87 (1.16) | 0.017 |
| Selenium (mcg) | 410.19 (78.99) | 494.54 (132.81) | 0.398 |
| Sodium (mg) | 15073.14 (7546.94) | 12668.31 (132.81) | 0.610 |
| Potassium (mg) | 36315.79 (14003.99) | 18735.86 (5889.65) | 0.116 |

AnimalP; animal protein diet, SoyP; soy-pea protein diet.

*Unpaired t-test p. Calculations include self-reported foods not provided by the study. Dietary intake data collected and analyzed using Nutrition Data System for Research software version 2019, developed by the Nutrition Coordinating Center (NCC), University of Minnesota, Minneapolis, MN. Results are mean, standard deviation.

**Supplementary Table 9.** Within-group Total Micronutrient Intake Over 7 Days Among Female Crohn's Disease Participants by Diet (SoyP vs AnimalP)

| | SoyP (n=8) | AnimalP (n=10) | P-value* |
|---|---|---|---|
| Daidzein (mg) | 232.75 (65.35) | 1.61 (4.73) | <0.000001 |
| Genistein (mg) | 289.16 (78.18) | 2.75 (6.64) | <0.000001 |
| Glycitein (mg) | 59.09 (14.07) | 0.35 (1.09) | <0.000001 |
| Coumestrol (mg) | 0.44 (0.34) | 0.07 (0.12) | 0.005 |
| Biochanin A (mg) | 0.06 (0.07) | 0.02 (0.02) | 0.175 |
| Formononetin (mg) | 0.10 (0.08) | 0.05 (0.04) | 0.087 |
| Tryptophan (g) | 5.68 (0.84) | 6.29 (1.60) | 0.344 |
| Threonine (g) | 17.11 (2.33) | 21.20 (5.04) | 0.050 |
| Isoleucine (g) | 19.21 (2.96) | 24.03 (6.33) | 0.065 |
| Leucine (g) | 19.21 (4.73) | 41.07 (10.17) | 0.00004 |
| Lysine (g) | 24.36 (3.20) | 36.24 (8.87) | 0.002 |
| Methionine (g) | 6.12 (0.94) | 36.24 (3.01) | <0.000001 |
| Cystine (g) | 6.06 (0.92) | 36.24 (1.81) | <0.000001 |
| Phenylalanine (g) | 22.69 (0.34) | 23.77 (6.38) | 0.640 |
| Tyrosine (g) | 15.33 (1.95) | 23.77 (5.12) | 0.0005 |
| Valine (g) | 21.39 (3.00) | 27.72 (7.45) | 0.039 |
| Arginine (g) | 39.69 (5.61) | 34.63 (10.10) | 0.224 |
| Histidine (g) | 11.83 (1.49) | 14.90 (3.73) | 0.044 |
| Alanine (g) | 20.15 (2.62) | 25.24 (6.30) | 0.049 |
| Aspartic Acid (g) | 54.42 (6.61) | 51.32 (13.24) | 0.555 |
| Glutamic Acid (g) | 94.96 (13.19) | 96.33 (23.77) | 0.886 |
| Glycine (g) | 21.63 (2.96) | 23.22 (6.50) | 0.531 |
| Proline (g) | 23.59 (3.53) | 30.78 (8.03) | 0.032 |
| Serine (g) | 24.05 (3.18) | 24.79 (6.48) | 0.772 |
| Vit A (IU) | 65145.57 (52687.07) | 66957.41 (29239.34) | 0.927 |
| Vit D (mcg) | 20.13 (8.04) | 16.22 (9.64) | 0.372 |
| Vit E (mg) | 99.70 (66.31) | 82.05 (50.63) | 0.530 |
| Vit K (mcg) | 1317.99 (542.40) | 1099.86 (602.34) | 0.437 |
| Vit C (mg) | 859.50 (574.11) | 789.11 (458.27) | 0.776 |
| Thiamin (mg) | 6.74 (1.61) | 4.52 (1.25) | 0.004 |
| Riboflavin (mg) | 8.52 (1.52) | 10.06 (2.80) | 0.184 |
| Niacin (mg) | 90.55 (16.92) | 124.14 (49.02) | 0.084 |
| Pantothenic Acid (mg) | 26.30 (9.68) | 34.94 (13.56) | 0.149 |
| Vit B-6 (mg) | 11.23 (2.59) | 13.26 (3.64) | 0.204 |
| Folate (mcg) | 3237.96 (929.21) | 1904.89 (696.20) | 0.003 |
| B- 12 (mcg) | 12.19 (7.59) | 23.08 (6.35) | 0.004 |
| Calcium (mg) | 4963.73 (867.80) | 5111.39 (2089.58) | 0.854 |
| Phosphorous (mg) | 7874.46 (1576.87) | 8437.84 (2631.57) | 0.602 |
| Magnesium (mg) | 3198.45 (514.45) | 2362.23 (717.41) | 0.014 |
| Iron (mg) | 104.39 (16.79) | 66.73 (20.76) | 0.001 |
| Zinc (mg) | 57.41 (10.67) | 74.52 (19.37) | 0.040 |
| Copper (mg) | 19.64 (4.29) | 11.46 (3.81) | 0.001 |
| Selenium (mcg) | 237.05 (89.17) | 520.06 (187.76) | 0.001 |
| Sodium (mg) | 13042.21 (5616.16) | 12510.68 (5090.05) | 0.836 |
| Potassium (mg) | 24195.35 (3951.84) | 20274.61 (5220.05) | 0.098 |

AnimalP; animal protein diet, SoyP; soy-pea protein diet.

*Unpaired t-test p. Calculations include self-reported foods not provided by the study. Dietary intake data collected and analyzed using Nutrition Data System for Research software version 2019, developed by the Nutrition Coordinating Center (NCC), University of Minnesota, Minneapolis, MN. Results are mean, standard deviation.

**Supplementary Table 10.** Within-group Total Micronutrient Intake Over 7 Days Among Female Healthy Control Participants by Diet (SoyP vs AnimalP)

| | SoyP (n=11) | AnimalP (n=12) | P-value* |
|---|---|---|---|
| Daidzein (mg) | 255.34 (86.37) | 0.15 (0.08) | <0.000001 |
| Genistein (mg) | 324.06 (111.44) | 0.62 (0.58) | <0.000001 |
| Glycitein (mg) | 54.27 (1.60) | 0.00 (0.00) | <0.000001 |
| Coumestrol (mg) | 0.34 (0.18) | 0.10 (0.14) | 0.002 |
| Biochanin A (mg) | 0.02 (0.01) | 1.19 (3.86) | 0.327 |
| Formononetin (mg) | 0.06 (0.04) | 0.06 (0.05) | 0.697 |
| Tryptophan (g) | 6.15 (2.38) | 7.21 (1.76) | 0.238 |
| Threonine (g) | 6.15 (6.89) | 23.45 (5.57) | 0.000001 |
| Isoleucine (g) | 21.02 (8.29) | 25.94 (6.72) | 0.131 |
| Leucine (g) | 35.84 (13.72) | 45.40 (11.18) | 0.080 |
| Lysine (g) | 26.61 (9.89) | 39.91 (9.93) | 0.004 |
| Methionine (g) | 6.60 (2.67) | 13.00 (3.15) | 0.000 |
| Cystine (g) | 6.60 (2.57) | 7.02 (1.59) | 0.635 |
| Phenylalanine (g) | 24.25 (9.48) | 26.01 (6.56) | 0.608 |
| Tyrosine (g) | 24.25 (6.15) | 20.81 (5.43) | 0.169 |
| Valine (g) | 23.15 (9.07) | 29.96 (7.43) | 0.061 |
| Arginine (g) | 40.61 (16.17) | 37.79 (9.92) | 0.615 |
| Histidine (g) | 12.60 (4.88) | 16.20 (4.13) | 0.069 |
| Alanine (g) | 21.49 (8.32) | 27.66 (6.32) | 0.057 |
| Aspartic Acid (g) | 57.79 (21.98) | 57.95 (13.84) | 0.984 |
| Glutamic Acid (g) | 99.31 (38.39) | 108.71 (27.43) | 0.503 |
| Glycine (g) | 22.64 (9.17) | 24.93 (6.30) | 0.490 |
| Proline (g) | 25.10 (9.48) | 34.33 (9.81) | 0.033 |
| Serine (g) | 25.44 (9.63) | 27.52 (6.74) | 0.551 |
| Vit A (IU) | 41217.13 (29235.27) | 55278.43 (27674.21) | 0.249 |
| Vit D (mcg) | 24.88 (11.11) | 42.35 (30.85) | 0.091 |
| Vit E (mg) | 112.45 (967.55) | 119.40 (32.59) | 0.980 |
| Vit K (mcg) | 1519.94 (914.91) | 1737.03 (738.63) | 0.536 |
| Vit C (mg) | 943.83 (530.10) | 1030.50 (425.22) | 0.668 |
| Thiamin (mg) | 7.55 (2.78) | 5.18 (1.68) | 0.021 |
| Riboflavin (mg) | 10.30 (3.60) | 12.08 (3.41) | 0.237 |
| Niacin (mg) | 83.14 (40.75) | 135.18 (44.60) | 0.008 |
| Pantothenic Acid (mg) | 30.15 (17.59) | 40.33 (7.65) | 0.082 |
| Vit B-6 (mg) | 11.77 (6.90) | 13.59 (2.88) | 0.411 |
| Folate (mcg) | 2993.92 (1203.95) | 2096.34 (606.86) | 0.033 |
| B- 12 (mcg) | 20.63 (12.43) | 26.27 (7.52) | 0.197 |
| Calcium (mg) | 5577.14 (2026.25) | 6189.01 (1603.56) | 0.429 |
| Phosphorous (mg) | 7980.69 (3484.80) | 9613.98 (2444.03) | 0.204 |
| Magnesium (mg) | 3208.01 (1296.63) | 2679.87 (680.01) | 0.229 |
| Iron (mg) | 115.64 (47.50) | 69.47 (16.04) | 0.004 |
| Zinc (mg) | 61.97 (32.24) | 76.76 (21.62) | 0.207 |
| Copper (mg) | 20.92 (8.24) | 13.00 (3.06) | 0.005 |
| Selenium (mcg) | 237.79 (116.99) | 537.01 (135.58) | 0.00001 |
| Sodium (mg) | 13167.80 (4756.00) | 14578.50 (3081.55) | 0.404 |
| Potassium (mg) | 25520.77 (9487.25) | 22884.26 (4832.22) | 0.404 |

AnimalP; animal protein diet, SoyP; soy-pea protein diet.

*Unpaired t-test p. Calculations include self-reported foods not provided by the study. Dietary intake data collected and analyzed using Nutrition Data System for Research software version 2019, developed by the Nutrition Coordinating Center (NCC), University of Minnesota, Minneapolis, MN. Results are mean, standard deviation.

**Supplementary Table 11.** Between-group Total Micronutrient Intake Over 7 Days Among Male Crohn's Disease and Healthy Control Participants Assigned the SoyP diet

| | Healthy Control (n=3) | Crohn's Disease (n=8) | P-value* |
|---|---|---|---|
| Daidzein (mg) | 331.28 (87.95) | 332.95 (101.10) | 0.981 |
| Genistein (mg) | 434.28 (115.04) | 428.83 (137.15) | 0.953 |
| Glycitein (mg) | 66.63 (17.11) | 79.88 (26.43) | 0.448 |
| Coumestrol (mg) | 0.26 (0.23) | 0.37 (0.38) | 0.669 |
| Biochanin A (mg) | 0.05 (0.01) | 0.74 (2.22) | 0.612 |
| Formononetin (mg) | 0.14 (0.02) | 0.07 (0.03) | 0.003 |
| Tryptophan (g) | 9.64 (2.86) | 7.73 (1.50) | 0.169 |
| Threonine (g) | 27.87 (8.54) | 22.65 (4.00) | 0.184 |
| Isoleucine (g) | 33.08 (10.12) | 26.38 (5.38) | 0.175 |
| Leucine (g) | 55.87 (17.30) | 44.74 (8.99) | 0.182 |
| Lysine (g) | 41.10 (12.59) | 33.97 (7.02) | 0.250 |
| Methionine (g) | 10.56 (3.10) | 8.18 (1.54) | 0.112 |
| Cystine (g) | 10.53 (3.09) | 7.98 (1.41) | 0.081 |
| Phenylalanine (g) | 38.12 (12.23) | 29.91 (6.00) | 0.156 |
| Tyrosine (g) | 25.11 (7.87) | 20.69 (4.10) | 0.240 |
| Valine (g) | 36.56 (10.99) | 28.67 (5.28) | 0.129 |
| Arginine (g) | 64.47 (20.33) | 49.87 (9.52) | 0.125 |
| Histidine (g) | 19.66 (6.32) | 15.58 (3.13) | 0.171 |
| Alanine (g) | 33.12 (10.61) | 26.27 (4.79) | 0.157 |
| Aspartic Acid (g) | 88.42 (29.18) | 70.80 (14.09) | 0.194 |
| Glutamic Acid (g) | 156.08 (49.12) | 121.34 (21.95) | 0.123 |
| Glycine (g) | 35.79 (11.99) | 26.99 (4.84) | 0.099 |
| Proline (g) | 38.69 (12.41) | 31.79 (6.65) | 0.249 |
| Serine (g) | 39.52 (12.28) | 31.95 (6.58) | 0.205 |
| Vit A (IU) | 46437.68 (17985.98) | 64184.54 (31563.76) | 0.391 |
| Vit D (mcg) | 32.09 (7.29) | 29.55 (8.54) | 0.661 |
| Vit E (mg) | 175.24 (116.50) | 68.64 (22.18) | 0.024 |
| Vit K (mcg) | 2078.73 (1405.88) | 1680.65 (622.16) | 0.512 |
| Vit C (mg) | 1520.68 (786.49) | 1215.37 (646.61) | 0.524 |
| Thiamin (mg) | 9.85 (3.01) | 8.92 (2.43) | 0.604 |
| Riboflavin (mg) | 13.23 (5.75) | 11.12 (2.09) | 0.368 |
| Niacin (mg) | 124.37 (50.67) | 94.01 (35.69) | 0.286 |
| Pantothenic Acid (mg) | 48.12 (20.12) | 28.95 (10.22) | 0.059 |
| Vit B-6 (mg) | 18.04 (6.39) | 12.37 (3.21) | 0.073 |
| Folate (mcg) | 4160.44 (1649.38) | 3710.79 (83.53) | 0.417 |
| B- 12 (mcg) | 24.93 (5.44) | 3710.79 (8.38) | <0.000001 |
| Calcium (mg) | 7095.85 (3053.84) | 6351.26 (1053.21) | 0.537 |
| Phosphorous (mg) | 13327.84 (3774.09) | 9198.60 (1622.75) | 0.026 |
| Magnesium (mg) | 4741.40 (1567.74) | 3430.84 (861.19) | 0.101 |
| Iron (mg) | 167.75 (54.40) | 135.19 (24.01) | 0.182 |
| Zinc (mg) | 94.80 (27.10) | 65.87 (15.39) | 0.048 |
| Copper (mg) | 32.94 (9.12) | 23.14 (22.74) | 0.498 |
| Selenium (mcg) | 410.19 (78.99) | 244.48 (71.65) | 0.009 |
| Sodium (mg) | 15073.14 (7546.94) | 16647.12 (9780.58) | 0.809 |
| Potassium (mg) | 36315.79 (14003.99) | 28267.74 (5368.04) | 0.177 |

AnimalP; animal protein diet, SoyP; soy-pea protein diet.

*Unpaired t-test p. Calculations include self-reported foods not provided by the study. Dietary intake data collected and analyzed using Nutrition Data System for Research software version 2019, developed by the Nutrition Coordinating Center (NCC), University of Minnesota, Minneapolis, MN. Results are mean, standard deviation.

**Supplementary Table 12**. Between-group Total Micronutrient Intake Over 7 Days Among Male Crohn's Disease and Healthy Control Participants Assigned the AnimalP diet

| | Healthy Control (n=3) | Crohn's Disease (n=5) | P-value* |
|---|---|---|---|
| Kcals | 1745.50 (212.60) | 1745.50 (212.60) | 0.558 |
| Fat (g) | 89.40 (14.90) | 89.40 (14.90) | 0.325 |
| Carb (g) | 171.20 (31.70) | 171.20 (31.70) | 0.543 |
| Pro (g) | 86.90 (13.10) | 86.90 (13.10) | 0.152 |
| Total Fiber | 137.80 (33.90) | 137.80 (33.90) | 0.601 |
| Daidzein (mg) | 0.12 (0.07) | 0.12 (0.07) | 0.350 |
| Genistein (mg) | 0.72 (0.68) | 0.72 (0.68) | 0.589 |
| Glycitein (mg) | 0.00 (0.00) | 0.00 (0.00) | 0.434 |
| Coumestrol (mg) | 0.04 (0.07) | 0.04 (0.07) | 0.323 |
| Biochanin A (mg) | 0.02 (0.02) | 0.02 (0.02) | 0.430 |
| Formononetin (mg) | 0.04 (0.02) | 0.04 (0.02) | 0.449 |
| Tryptophan (g) | 6.39 (1.25) | 6.39 (1.25) | 0.241 |
| Threonine (g) | 21.05 (4.04) | 21.05 (4.04) | 0.121 |
| Isoleucine (g) | 23.83 (4.67) | 23.83 (4.67) | 0.148 |
| Leucine (g) | 40.58 (7.25) | 40.58 (7.25) | 0.200 |
| Lysine (g) | 35.96 (7.00) | 35.96 (7.00) | 0.202 |
| Methionine (g) | 11.63 (2.39) | 11.63 (2.39) | 0.127 |
| Cystine (g) | 6.71 (0.90) | 6.71 (0.90) | 0.264 |
| Phenylalanine (g) | 23.28 (4.56) | 23.28 (4.56) | 0.226 |
| Tyrosine (g) | 18.10 (4.27) | 18.10 (4.27) | 0.120 |
| Valine (g) | 27.03 (5.28) | 27.03 (5.28) | 0.204 |
| Arginine (g) | 36.42 (4.42) | 36.42 (4.42) | 0.560 |
| Histidine (g) | 14.52 (2.36) | 14.52 (2.36) | 0.222 |
| Alanine (g) | 25.24 (4.15) | 25.24 (4.15) | 0.219 |
| Aspartic Acid (g) | 52.08 (10.84) | 52.08 (10.84) | 0.337 |
| Glutamic Acid (g) | 97.25 (18.35) | 97.25 (18.35) | 0.340 |
| Glycine (g) | 22.99 (2.68) | 22.99 (2.68) | 0.302 |
| Proline (g) | 27.49 (7.16) | 27.49 (7.16) | 0.133 |
| Serine (g) | 24.26 (4.65) | 24.26 (4.65) | 0.246 |
| Vit A (IU) | 25606.07 (20788.15) | 25606.07 (20788.15) | 0.005 |
| Vit D (mcg) | 39.33 (51.87) | 39.33 (51.87) | 0.545 |
| Vit E (mg) | 73.31 (18.16) | 73.31 (18.16) | 0.812 |
| Vit K (mcg) | 1147.01 (1114.83) | 1147.01 (1114.83) | 0.421 |
| Vit C (mg) | 873.48 (873.09) | 873.48 (873.09) | 0.896 |
| Thiamin (mg) | 4.61 (0.77) | 4.61 (0.77) | 0.589 |
| Riboflavin (mg) | 9.65 (3.67) | 9.65 (3.67) | 0.335 |
| Niacin (mg) | 127.40 (37.03) | 127.40 (37.03) | 0.436 |
| Pantothenic Acid (mg) | 33.16 (12.99) | 33.16 (12.99) | 0.407 |
| Vit B-6 (mg) | 12.27 (3.86) | 12.27 (3.86) | 0.490 |
| Folate (mcg) | 1520.37 (1003.23) | 1520.37 (1003.23) | 0.283 |
| B- 12 (mcg) | 20.10 (7.79) | 20.10 (7.79) | 0.098 |
| Calcium (mg) | 4054.93 (1727.81) | 4054.93 (1727.81) | 0.044 |
| Phosphorous (mg) | 8144.98 (1688.50) | 8144.98 (1688.50) | 0.409 |
| Magnesium (mg) | 2496.95 (237.06) | 2496.95 (237.06) | 0.997 |
| Iron (mg) | 63.10 (5.24) | 63.10 (5.24) | 0.443 |
| Zinc (mg) | 65.57 (11.15) | 65.57 (11.15) | 0.337 |
| Copper (mg) | 11.87 (1.16) | 11.87 (1.16) | 0.978 |
| Selenium (mcg) | 494.54 (132.81) | 494.54 (132.81) | 0.308 |
| Sodium (mg) | 12668.31 (132.81) | 12668.31 (132.81) | 0.190 |
| Potassium (mg) | 18735.86 (5889.65) | 18735.86 (5889.65) | 0.495 |

AnimalP; animal protein diet, SoyP; soy-pea protein diet.

*Unpaired t-test p. Calculations include self-reported foods not provided by the study. Dietary intake data collected and analyzed using Nutrition Data System for Research software version 2019, developed by the Nutrition Coordinating Center (NCC), University of Minnesota, Minneapolis, MN. Results are mean, standard deviation.

**Supplementary Table 13.** Between-group Total Micronutrient Intake Over 7 Days Among Female Crohn's Disease and Healthy Control Participants Assigned the SoyP diet

| | Healthy Control (n=11) | Crohn's Disease (n=8) | P-value* |
|---|---|---|---|
| Daidzein (mg) | 255.34 (86.37) | 232.75 (65.35) | 0.543 |
| Genistein (mg) | 324.06 (111.44) | 289.16 (78.18) | 0.459 |
| Glycitein (mg) | 54.27 (1.60) | 59.09 (14.07) | 0.271 |
| Coumestrol (mg) | 0.34 (0.18) | 0.44 (0.34) | 0.420 |
| Biochanin A (mg) | 0.02 (0.01) | 0.06 (0.07) | 0.115 |
| Formononetin (mg) | 0.06 (0.04) | 0.10 (0.08) | 0.212 |
| Tryptophan (g) | 6.15 (2.38) | 5.68 (0.84) | 0.600 |
| Threonine (g) | 6.15 (6.89) | 17.11 (2.33) | 0.0005 |
| Isoleucine (g) | 21.02 (8.29) | 19.21 (2.96) | 0.564 |
| Leucine (g) | 35.84 (13.72) | 19.21 (4.73) | 0.005 |
| Lysine (g) | 26.61 (9.89) | 24.36 (3.20) | 0.546 |
| Methionine (g) | 6.60 (2.67) | 6.12 (0.94) | 0.636 |
| Cystine (g) | 6.60 (2.57) | 6.06 (0.92) | 0.580 |
| Phenylalanine (g) | 24.25 (9.48) | 22.69 (0.34) | 0.650 |
| Tyrosine (g) | 24.25 (6.15) | 15.33 (1.95) | 0.001 |
| Valine (g) | 23.15 (9.07) | 21.39 (3.00) | 0.606 |
| Arginine (g) | 40.61 (16.17) | 39.69 (5.61) | 0.880 |
| Histidine (g) | 12.60 (4.88) | 11.83 (1.49) | 0.673 |
| Alanine (g) | 21.49 (8.32) | 20.15 (2.62) | 0.669 |
| Aspartic Acid (g) | 57.79 (21.98) | 54.42 (6.61) | 0.682 |
| Glutamic Acid (g) | 99.31 (38.39) | 94.96 (13.19) | 0.764 |
| Glycine (g) | 22.64 (9.17) | 21.63 (2.96) | 0.767 |
| Proline (g) | 25.10 (9.48) | 23.59 (3.53) | 0.673 |
| Serine (g) | 25.44 (9.63) | 24.05 (3.18) | 0.701 |
| Vit A (IU) | 41217.13 (29235.27) | 65145.57 (52687.07) | 0.221 |
| Vit D (mcg) | 24.88 (11.11) | 20.13 (8.04) | 0.319 |
| Vit E (mg) | 112.45 (967.55) | 99.70 (66.31) | 0.971 |
| Vit K (mcg) | 1519.94 (914.91) | 1317.99 (542.40) | 0.586 |
| Vit C (mg) | 943.83 (530.10) | 859.50 (574.11) | 0.745 |
| Thiamin (mg) | 7.55 (2.78) | 6.74 (1.61) | 0.472 |
| Riboflavin (mg) | 10.30 (3.60) | 8.52 (1.52) | 0.208 |
| Niacin (mg) | 83.14 (40.75) | 90.55 (16.92) | 0.636 |
| Pantothenic Acid (mg) | 30.15 (17.59) | 26.30 (9.68) | 0.584 |
| Vit B-6 (mg) | 11.77 (6.90) | 11.23 (2.59) | 0.838 |
| Folate (mcg) | 2993.92 (1203.95) | 3237.96 (929.21) | 0.639 |
| B- 12 (mcg) | 20.63 (12.43) | 12.19 (7.59) | 0.108 |
| Calcium (mg) | 5577.14 (2026.25) | 4963.73 (867.80) | 0.435 |
| Phosphorous (mg) | 7980.69 (3484.80) | 7874.46 (1576.87) | 0.937 |
| Magnesium (mg) | 3208.01 (1296.63) | 3198.45 (514.45) | 0.985 |
| Iron (mg) | 115.64 (47.50) | 104.39 (16.79) | 0.532 |
| Zinc (mg) | 61.97 (32.24) | 57.41 (10.67) | 0.707 |
| Copper (mg) | 20.92 (8.24) | 19.64 (4.29) | 0.696 |
| Selenium (mcg) | 237.79 (116.99) | 237.05 (89.17) | 0.988 |
| Sodium (mg) | 13167.80 (4756.00) | 13042.21 (5616.16) | 0.959 |
| Potassium (mg) | 25520.77 (9487.25) | 24195.35 (3951.84) | 0.716 |

AnimalP; animal protein diet, SoyP; soy-pea protein diet.

*Unpaired t-test p. Calculations include self-reported foods not provided by the study. Dietary intake data collected and analyzed using Nutrition Data System for Research software version 2019, developed by the Nutrition Coordinating Center (NCC), University of Minnesota, Minneapolis, MN. Results are mean, standard deviation.

**Supplementary Table 14.** Between-group Total Micronutrient Intake Over 7 Days Among Female Crohn's Disease and Healthy Control Participants Assigned the AnimalP diet

| | Healthy Control (n=12) | Crohn's Disease (n=10) | P-value* |
|---|---|---|---|
| Daidzein (mg) | 0.15 (0.08) | 1.61 (4.73) | 0.294 |
| Genistein (mg) | 0.62 (0.58) | 2.75 (6.64) | 0.279 |
| Glycitein (mg) | 0.00 (0.00) | 0.35 (1.09) | 0.284 |
| Coumestrol (mg) | 0.10 (0.14) | 0.07 (0.12) | 0.564 |
| Biochanin A (mg) | 1.19 (3.86) | 0.02 (0.02) | 0.352 |
| Formononetin (mg) | 0.06 (0.05) | 0.05 (0.04) | 0.632 |
| Tryptophan (g) | 7.21 (1.76) | 6.29 (1.60) | 0.222 |
| Threonine (g) | 23.45 (5.57) | 21.20 (5.04) | 0.335 |
| Isoleucine (g) | 25.94 (6.72) | 24.03 (6.33) | 0.503 |
| Leucine (g) | 45.40 (11.18) | 41.07 (10.17) | 0.358 |
| Lysine (g) | 39.91 (9.93) | 36.24 (8.87) | 0.376 |
| Methionine (g) | 13.00 (3.15) | 36.24 (3.01) | <0.000001 |
| Cystine (g) | 7.02 (1.59) | 36.24 (1.81) | <0.000001 |
| Phenylalanine (g) | 26.01 (6.56) | 23.77 (6.38) | 0.429 |
| Tyrosine (g) | 20.81 (5.43) | 23.77 (5.12) | 0.206 |
| Valine (g) | 29.96 (7.43) | 27.72 (7.45) | 0.489 |
| Arginine (g) | 37.79 (9.92) | 34.63 (10.10) | 0.470 |
| Histidine (g) | 16.20 (4.13) | 14.90 (3.73) | 0.452 |
| Alanine (g) | 27.66 (6.32) | 25.24 (6.30) | 0.381 |
| Aspartic Acid (g) | 57.95 (13.84) | 51.32 (13.24) | 0.268 |
| Glutamic Acid (g) | 108.71 (27.43) | 96.33 (23.77) | 0.276 |
| Glycine (g) | 24.93 (6.30) | 23.22 (6.50) | 0.538 |
| Proline (g) | 34.33 (9.81) | 30.78 (8.03) | 0.371 |
| Serine (g) | 27.52 (6.74) | 24.79 (6.48) | 0.346 |
| Vit A (IU) | 55278.43 (27674.21) | 66957.41 (29239.34) | 0.348 |
| Vit D (mcg) | 42.35 (30.85) | 16.22 (9.64) | 0.018 |
| Vit E (mg) | 119.40 (32.59) | 82.05 (50.63) | 0.049 |
| Vit K (mcg) | 1737.03 (738.63) | 1099.86 (602.34) | 0.041 |
| Vit C (mg) | 1030.50 (425.22) | 789.11 (458.27) | 0.215 |
| Thiamin (mg) | 5.18 (1.68) | 4.52 (1.25) | 0.312 |
| Riboflavin (mg) | 12.08 (3.41) | 10.06 (2.80) | 0.149 |
| Niacin (mg) | 135.18 (44.60) | 124.14 (49.02) | 0.587 |
| Pantothenic Acid (mg) | 40.33 (7.65) | 34.94 (13.56) | 0.254 |
| Vit B-6 (mg) | 13.59 (2.88) | 13.26 (3.64) | 0.815 |
| Folate (mcg) | 2096.34 (606.86) | 1904.89 (696.20) | 0.498 |
| B- 12 (mcg) | 26.27 (7.52) | 23.08 (6.35) | 0.301 |
| Calcium (mg) | 6189.01 (1603.56) | 5111.39 (2089.58) | 0.186 |
| Phosphorous (mg) | 9613.98 (2444.03) | 8437.84 (2631.57) | 0.291 |
| Magnesium (mg) | 2679.87 (680.01) | 2362.23 (717.41) | 0.300 |
| Iron (mg) | 69.47 (16.04) | 66.73 (20.76) | 0.730 |
| Zinc (mg) | 76.76 (21.62) | 74.52 (19.37) | 0.802 |
| Copper (mg) | 13.00 (3.06) | 11.46 (3.81) | 0.307 |
| Selenium (mcg) | 537.01 (135.58) | 520.06 (187.76) | 0.808 |
| Sodium (mg) | 14578.50 (3081.55) | 12510.68 (5090.05) | 0.254 |
| Potassium (mg) | 22884.26 (4832.22) | 20274.61 (5220.05) | 0.238 |

AnimalP; animal protein diet, SoyP; soy-pea protein diet.

*Unpaired t-test p. Calculations include self-reported foods not provided by the study. Dietary intake data collected and analyzed using Nutrition Data System for Research software version 2019, developed by the Nutrition Coordinating Center (NCC), University of Minnesota, Minneapolis, MN. Results are mean, standard deviation.

### D. Supplementary Figure

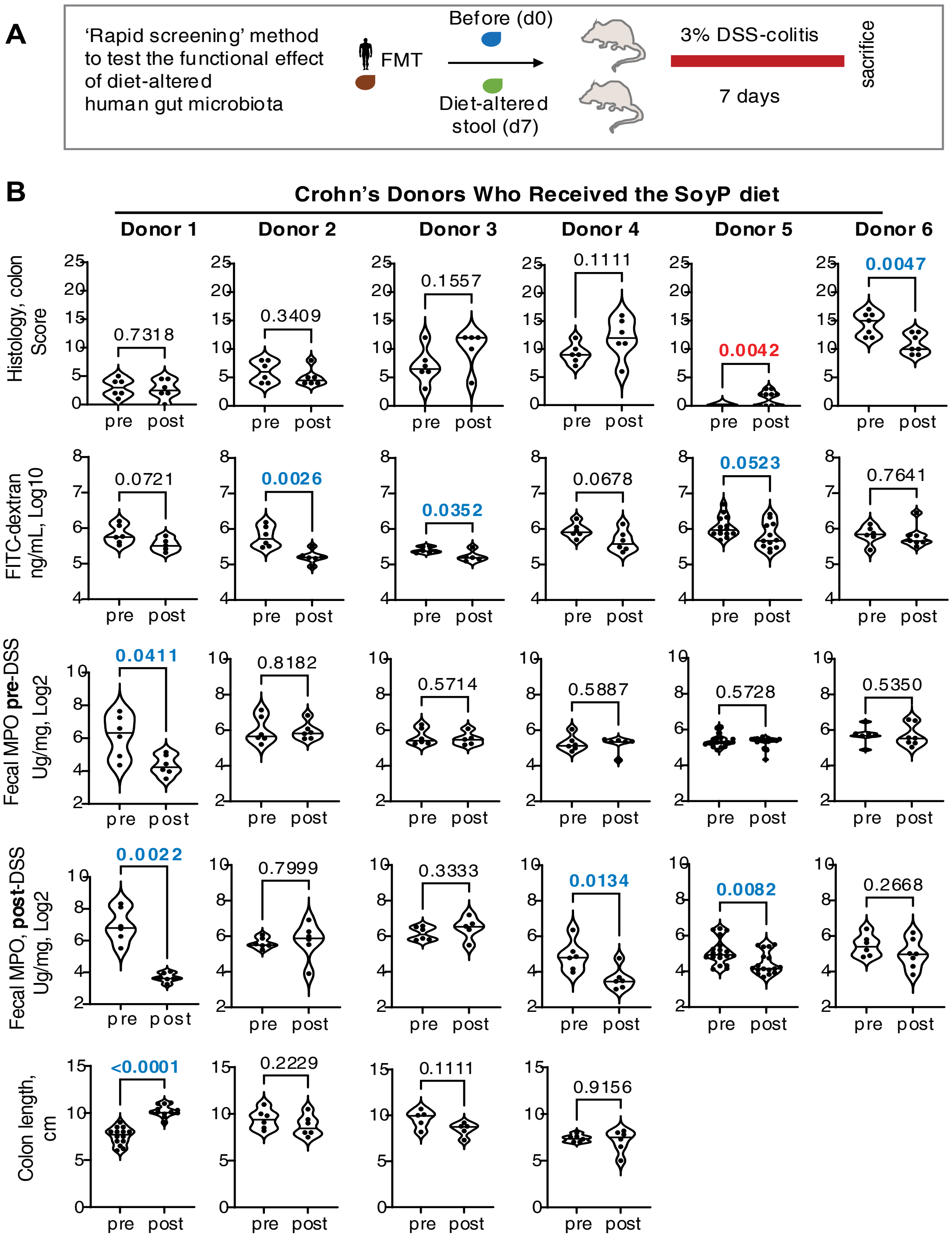


**Supplementary Figure 2. Fecal microbiota transplantation experiments using baseline ("pre") and diet-altered ("post") stool collected from Crohn's disease participants assigned to the SoyP-diet intervention. A)** Study Design for Fecal Microbiota Transplantation Experiments. Germ free SAMP mice transplanted with baseline ('pre'; d0) or diet-altered stool ('post'; d7) and then treated with 3% DSS for 7 days (n=6 mice/group). **B)** Colitis severity indices including histology, gut permeability (FITC-dextran), fecal myeloperoxidase (MPO), and colon length. Gut permeability and fecal MPO were measured *in vivo* before and after DSS induction. Colon length data missing for donor 5 and 6. Significant p-values where an improvement was observed indicated in bold blue font, and p-values where worsening was observed indicated in bold red font.
